# A Common Missense Variant, W335S, in β2-Glycoprotein I (APOH) is Associated with Increased Autoantibody Levels but Reduced Venous Thromboembolism Risk

**DOI:** 10.64898/2026.03.04.26347632

**Authors:** Christophe Lalaurie, Lili Liu, Atlas Khan, Chen Wang, Stephen S. Rich, R. Graham Barr, Elana J. Bernstein, Krzysztof Kiryluk, Thomas McDonnell, Yiming Luo

**Affiliations:** Aging, Rheumatology & Regenerative Medicine, Division of Medicine, University College London, London, UK; Department of Internal Medicine (Nephrology) and Department of Computational Medicine and Bioinformatics, University of Michigan, Ann Arbor, MI, USA; Division of Nephrology, Department of Medicine, Columbia University Irving Medical Center, New York, NY, USA; Department of Genome Sciences, University of Virginia, Charlottesville, VA, USA; Division of General Medicine, Department of Medicine, Columbia University Irving Medical Center, New York, NY, USA; Division of Rheumatology and Clinical Immunology, Department of Medicine, Columbia University Irving Medical Center, New York, NY, USA

## Abstract

Anti-β2-glycoprotein I (anti-β2GPI) antibodies are central to the pathogenesis of antiphospholipid syndrome (APS), an autoimmune disease characterized by a strong predisposition to venous thromboembolism (VTE). In this study, we conducted a multi-ancestry genome-wide association study (GWAS) of quantitative total anti-β2GPI levels in 5,969 participants enrolled in the Multi-Ethnic Study of Atherosclerosis (MESA) and identified a genome-wide significant association at the *APOH* locus. Paradoxically, genetically determined increases in anti-β2GPI levels at this locus were associated with lower VTE risk. Fine-mapping and functional genomics prioritized the missense variant rs1801690 (W335S) in β2GPI (apolipoprotein H, [APOH]) as the most likely causal variant. This variant has an allele frequency of 5-6% in European and East Asian ancestries but only 1% in African ancestries. Integrating prior experimental studies, molecular dynamics simulations and structure-based epitope prediction, we propose a dual-effect mechanism whereby W335S reduces thrombotic risk by disrupting phospholipid binding in Domain V, yet increases autoantibody production through conformational changes that enhance epitope exposure in Domains I and II. These findings mechanistically uncouple autoantibody formation from thrombotic risk in carriers of the W335S variant, and suggest that *APOH* genotype may represent a clinically relevant genetic biomarker with potential utility for thrombotic risk stratification in anti-β2GPI-positive individuals.

## Introduction

Anti-β2-glycoprotein I (anti-β2GPI) antibodies are key mediators of antiphospholipid syndrome (APS), a systemic autoimmune disorder characterized by a strong prothrombotic tendency leading to recurrent thrombosis and obstetric complications.^1^ β2GPI, also known as apolipoprotein H (APOH), is the autoantigen targeted by anti-β2GPI antibodies and is encoded by the *APOH* gene. As a plasma glycoprotein synthesized and secreted by the liver, β2GPI exhibits primarily anti-coagulant properties and binds to anionic phospholipids.^2^

The mechanism by which pathological anti-β2GPI antibodies induce thrombosis is closely linked to the structural biology of β2GPI.^3,4^ β2GPI is composed of five domains (Domain I [DI] to Domain V [DV]) with pathological anti-β2GPI antibodies recognizing epitopes within DI and DII, while DV mediates β2GPI binding to anionic phospholipids on cell membranes.^5^ Importantly, these antibodies bind to DI and induce thrombosis only when β2GPI binds to phospholipids via DV. Key epitope residues within DI specifically associated with thrombotic APS have been mapped to short peptide segments 29-36, 35-43, and 69–77, as well as residues 58–62 and 84–91, which represent the strongest antibody-binding sites within this domain.^6–9^ The autoantibody binding capacity of DI can be influenced not only by local conformational changes within DI itself but also by alterations in DV, such as plasmin-mediated cleavage, which can trigger a global conformational change in β2GPI.^10,11^

The precise mechanism underlying this phospholipid-dependent antibody binding to DI remains debated. A 2010 study hypothesized that β2GPI circulates in plasma as a compact, circular (O-Shape) conformation, masking cryptic DI epitopes; binding of DV to negatively charged phospholipids triggers a transition to an elongated J-Shape, exposing these sites.^12^ This model was later challenged in 2023 by Kumar et al., who did not observe evidence for an O-Shape conformation using advanced structural methods and instead suggested that phospholipid engagement by the J-shaped form elevates local β2GPI concentration, which enhances antibody recognition and downstream signaling.^13^ Conversely, we recently demonstrated through molecular dynamics simulations that a theoretical O-Shape can spontaneously unfold, with DV detaching from DI to form a J-shape and two subsequent S-Shape conformations, unmasking anti-β2GPI epitopes hidden in the O conformation at the residue-level.^11^ Our results potentially explain the failure to detect the O-Shape experimentally, as reported by Kumar et al., due to its inherent instability in solution.

A prior genome-wide association study (GWAS) in Europeans identified an *APOH* locus linked to positive anti-β2GPI antibody IgG.^14^ However, its impact on venous thromboembolism (VTE) risk and the underlying causal variant remain unclear. We therefore conducted a multi-ancestry GWAS meta-analysis of quantitative total anti-β2GPI antibody levels in a large population-based cohort and replicated the *APOH* locus association. Unexpectedly, we found that genetically determined increases in anti-β2GPI antibody levels due to *APOH* polymorphisms were paradoxically associated with a lower risk of VTE. We then identified a missense variant in *APOH* (β2GPI), W335S, as the likely causal variant mediating this paradoxical relationship. To further investigate the underlying mechanism, we performed molecular dynamics simulations starting from both O- and J-Shape β2GPI models. These analyses revealed a potential structural basis for how the W335S substitution may enhance autoantibody production while simultaneously reducing thrombotic risk.

## Results

### Genome-wide association studies

We included a total of 5,969 participants with both phenotype and high-quality genotype data in the GWAS (2,354 of European [EUR], 1,572 of African [AFR], 1,294 of Admixed American [AMR], and 749 of East Asian [EAS] ancestry). In the primary analysis of total anti-β2GPI antibody levels, the multi-ancestry GWAS meta-analysis identified a genome-wide significant association at the *APOH* locus, with the lead variant rs1801690-G (β = 0.21, p = 1.08 × 10⁻¹). There was no evidence of genomic inflation (λ = 0.99) (Figure 1, Supplementary Table S1).

**Figure 1.**
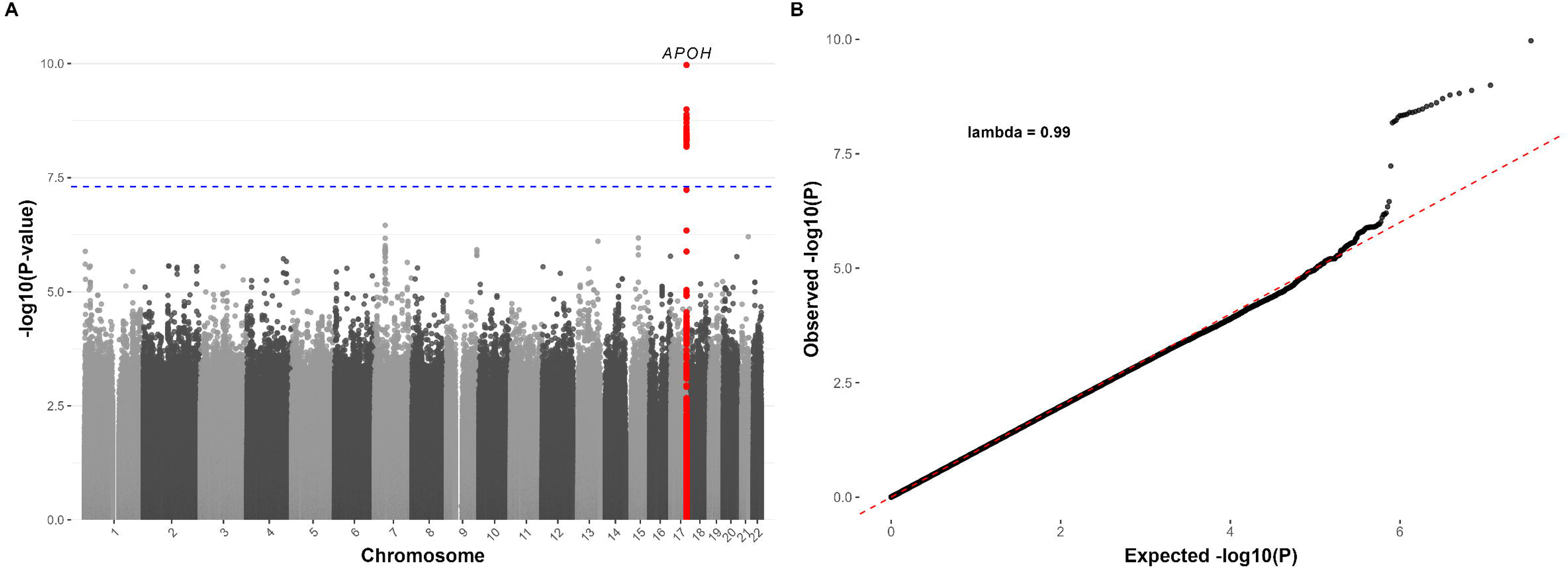
Manhattan and QQ plots of GWAS for total anti-β2GPI antibody levels.

In secondary analyses of anti-β2GPI antibody isotypes (IgM, IgG, and IgA) across different cutoffs, the *APOH* locus reached genome-wide significance for anti-β2GPI IgM at the 90th percentile (p = 8.44 × 10⁻¹), 95th percentile (p = 7.24 × 10⁻¹), and manufacturer’s cutoff (p = 5.96 × 10⁻¹), and for anti-β2GPI IgG at the 90th percentile (p = 3.34 × ⁻¹) (Supplementary Table S2, Supplementary Figure S2).

### Statistical fine-mapping

We used SuSiE as the primary approach for fine-mapping. At the *APOH* locus, we identified a single 95% credible set containing 18 variants. The variant with the highest posterior inclusion probability (PIP) was the GWAS lead variant rs1801690 (PIP = 0.49, Figure 2). All other variants in the credible set had substantially lower PIPs (all <0.07, Table 1 and Supplementary Table S3).

**Figure 2.**
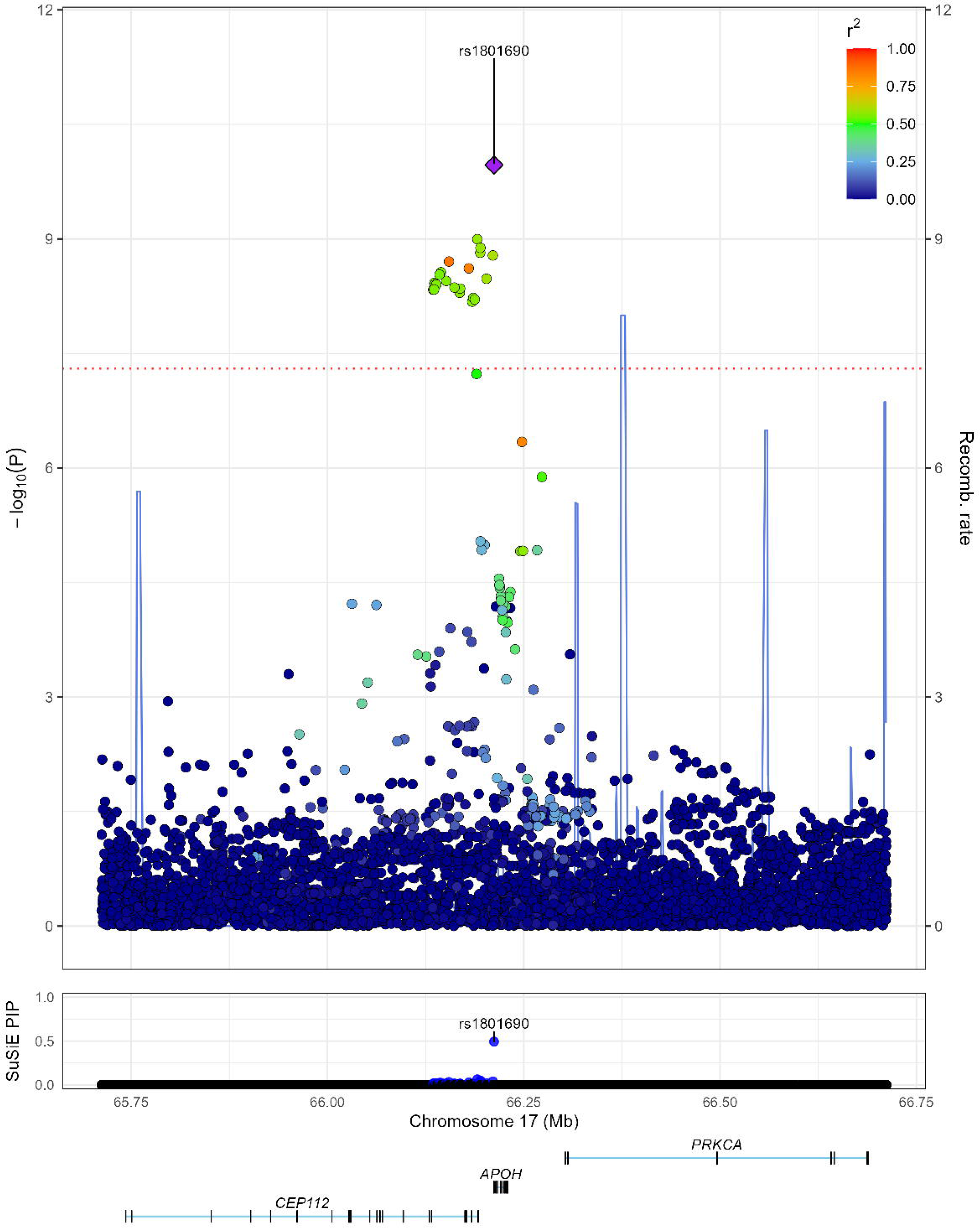
Regional plot of the *APOH* region with corresponding SuSiE posterior inclusion probabilities (PIP). The GWAS lead variant, rs1801690, was assigned the highest PIP (0.49) by SuSiE.

**Table 1.** Candidate causal variants in the 95 % credible set identified by SuSiE. Among these variants, rs1801690 had the highest posterior inclusion probability (PIP = 0.49) and is located in the exonic region of the *APOH* gene, whereas all remaining variants are located in nearby non-coding regions.

| rsID | CHR | BP | A1 | A2 | EAF | PIP | Annotation | Nearest Gene |
| --- | --- | --- | --- | --- | --- | --- | --- | --- |
| rs1801690 | 17 | 66212167 | G | C | 0.05 | 0.49 | exonic | *APOH* |
| rs9902706 | 17 | 66190870 | T | C | 0.06 | 0.06 | intronic | *CEP112* |
| rs9906486 | 17 | 66194802 | T | G | 0.06 | 0.05 | intergenic | *CEP112* |
| rs9905408 | 17 | 66194566 | C | A | 0.06 | 0.04 | intergenic | *CEP112* |
| rs7211380 | 17 | 66210650 | G | A | 0.06 | 0.04 | intergenic | *APOH* |
| rs74934196 | 17 | 66154754 | A | AAC | 0.05 | 0.03 | intronic | *CEP112* |
| rs74531840 | 17 | 66179997 | A | T | 0.05 | 0.03 | intronic | *CEP112* |
| rs76375367 | 17 | 66142666 | A | G | 0.06 | 0.02 | intronic | *CEP112* |
| rs55657678 | 17 | 66144550 | A | G | 0.06 | 0.02 | intronic | *CEP112* |
| rs11651658 | 17 | 66202522 | C | T | 0.06 | 0.02 | intergenic | *APOH* |
| rs73992250 | 17 | 66136049 | G | A | 0.06 | 0.02 | intronic | *CEP112* |
| rs7216660 | 17 | 66151258 | T | C | 0.06 | 0.02 | intronic | CEP112 |
| rs9891968 | 17 | 66136728 | A | G | 0.06 | 0.02 | intronic | CEP112 |
| rs9908597 | 17 | 66138565 | G | T | 0.06 | 0.02 | intronic | CEP112 |
| rs9895407 | 17 | 66161737 | C | T | 0.06 | 0.02 | intronic | CEP112 |
| rs77620153 | 17 | 66134676 | A | G | 0.06 | 0.02 | intronic | CEP112 |
| rs73992258 | 17 | 66168965 | C | A | 0.06 | 0.02 | intronic | CEP112 |
| rs9911603 | 17 | 66135784 | T | C | 0.06 | 0.02 | intronic | CEP112 |

CHR: chromosome; BP: base pair position; A1: effect allele; A2: non-effect allele; EAF: effect allele frequency; PIP : posterior inclusion probability.

In secondary multi-ancestry fine-mapping using MESuSiE, we again identified a single 95% credible set, which contained 21 variants and classified the underlying causal effect as shared across EUR, AMR, and EAS. Within this credible set, rs1801690 again had the highest PIP (0.23), followed by rs9906486 (PIP = 0.12) and rs9905408 (PIP = 0.12) (Supplementary Table S4). Regional plots for each ancestry group are shown in Supplementary Figure S3. The distribution of expected causal signals across ancestry configurations from MESuSiE is shown in Supplementary Figure S4.

The variant rs1801690 had a minor allele frequency (MAF) of 5.29% in EUR, 0.99% in AFR, 1.93% in AMR, and 6.14% in EAS. In the SuSiE-derived credible set, rs1801690 was in moderate to strong LD with the other 17 variants in EUR (median r² = 0.91), AMR (median r² = 0.62), and EAS (median r² = 0.74), but not in AFR (median r² = 0.12). The LD structure of the 18 variants in the multi-ancestry meta-analysis and within each ancestry group is presented in Supplementary Figure S5.

### Mendelian randomization and colocalization

We next performed Mendelian randomization (MR) and colocalization to examine the relationship between total anti-β2GPI antibody levels and VTE at the *APOH* locus, using GWAS summary statistics for VTE from a recent large meta-analysis.^15^ Due to extreme regional LD in the outcome (VTE) dataset resulting in a near-singular correlation structure among variants, we were unable to derive stable estimates using Generalized Inverse-Variance Weighted (GIVW) or principal component generalized method of moments (PC-GMM) methods. We therefore applied single-variant MR using the Wald ratio, followed by formal colocalization to assess whether the two traits shared a causal signal.^16^

The lead variant rs1801690 was a strong instrument for total anti-β2GPI antibody levels (F-statistic = 42). Unexpectedly, genetically determined anti-β2GPI antibody levels at the *APOH* locus were negatively associated with VTE (Beta = −0.25, p = 3.95 x 10^-6^). At the *APOH* locus, genetic association signals for anti-β2GPI antibody levels colocalized with VTE (PP4 = 0.97, Figure 3), with rs1801690 inferred as the lead candidate variant for the shared causal signal (per-variant PP4 = 0.74).

**Figure 3.**
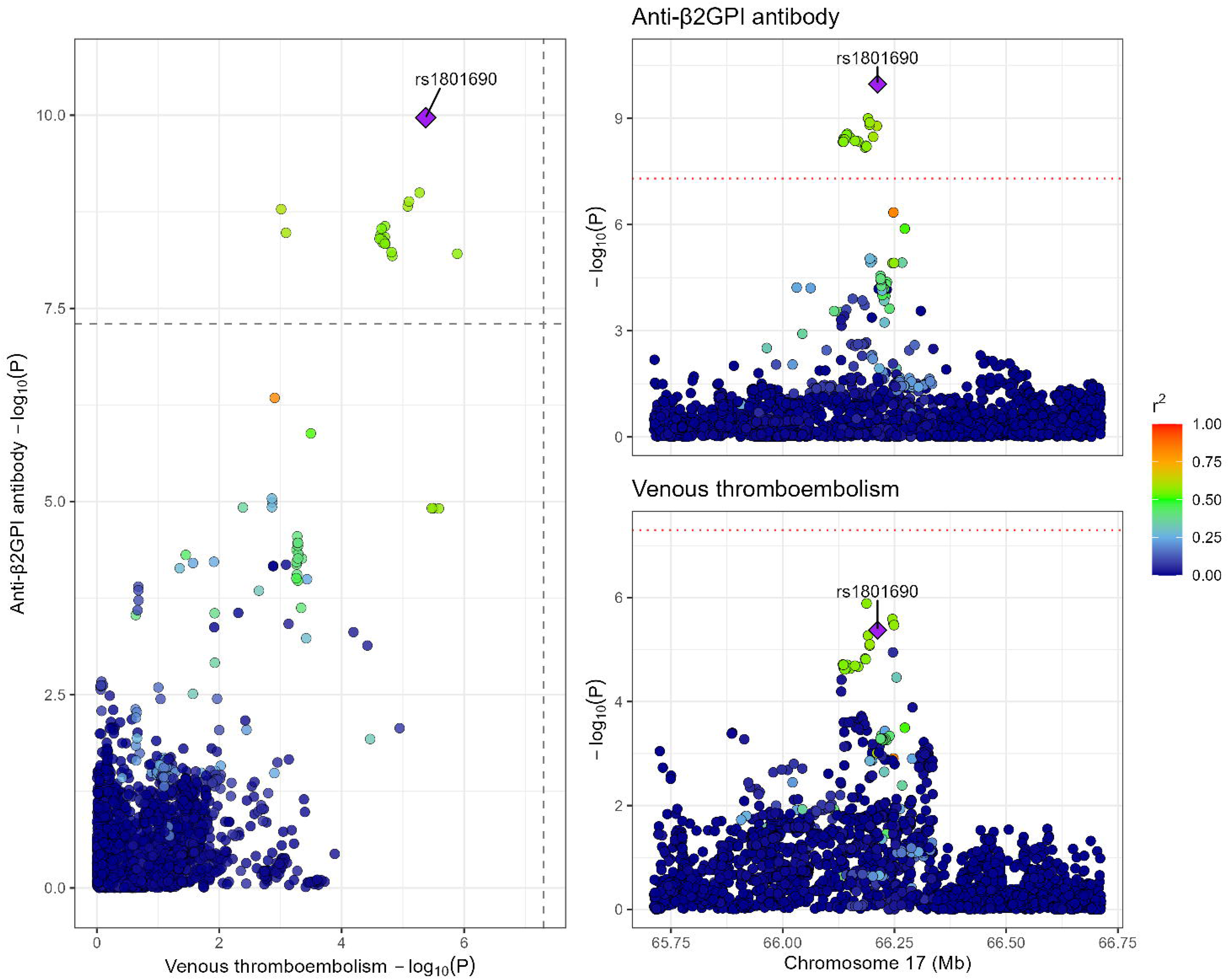
Colocalization plot of the *APOH* region between total anti-β2GPI antibody levels and venous thromboembolism (VTE). The genetic association signals colocalized with high confidence (PP4 = 0.97), with rs1801690 inferred as the lead candidate shared causal variant (per-variant PP4 = 0.74).

The finding was consistent across MR and colocalization analyses using positive anti-β2GPI IgG (90th percentile cutoff) and anti-β2GPI IgM (both manufacturer’s cutoff and 90th percentile cutoff, Supplementary Note). Consistent results were also observed with MR using positive anti-β2GPI IgG (99th percentile cutoff) from an independent GWAS of 4,163 German individuals (Supplementary Note).^14^

### Causal variant prioritization

Given the well-established role of anti-β2GPI antibodies in promoting thrombosis, the paradoxical association we observed, in which genetically determined higher anti-β2GPI antibody levels were associated with reduced VTE risk, likely reflects a violation of MR assumptions rather than a true protective effect of wild-type (WT) antibodies. We hypothesized that this relationship is instead mediated by a downstream effect of the causal variant within the *APOH* locus through alterations in β2GPI, which is both the cis-regulated product and the autoantigen of anti-β2GPI antibodies. The causal variant could either reside in a non-coding region, leading to changes in β2GPI expression levels, or in a protein-coding region, resulting in structural alterations that affect β2GPI function and immunogenicity.

To identify biologically plausible causal variants underlying the paradoxical association between higher anti-β2GPI antibody levels and reduced VTE risk, we applied a predefined variant prioritization framework to variants within the 95% credible set. We focused on non-coding variants with predicted regulatory effects on β2GPI (APOH) expression and non-synonymous protein-coding variants. We also hypothesized that increased β2GPI (APOH) expression could represent a regulatory mechanism linking higher autoantibody levels to reduced thrombosis.

Of the 18 variants in the 95% credible set, 17 were located in non-coding regions. The variant rs9902706 overlapped with an H3K4me1 GappedPeak but not a NarrowPeak. The variant rs7211380 overlapped with a H3K27ac GappedPeak but not a NarrowPeak. None of the non-coding variants in the credible set overlapped with liver cCREs from ENCODE or were predicted by ENCODE-rE2G to regulate β2GPI (APOH) expression in liver-related tissues or cells (Table 2). None of the 17 variants was associated with increased β2GPI (APOH) at either the liver mRNA or plasma protein level. In fact, all were associated with reduced plasma β2GPI levels (Table 2). We then tested colocalization of genetic association signals between total anti-β2GPI antibody levels from this study and (1) liver β2GPI (APOH) mRNA expression from GTEx; and (2) plasma β2GPI (APOH) protein levels from the UK Biobank. No evidence of colocalization was observed between total anti-β2GPI antibody levels and liver β2GPI (APOH) mRNA expression (PP4 < 0.01), or between total anti-β2GPI antibody levels and plasma β2GPI (APOH) protein levels (maximum PP4 = 0.11 across all SuSiE component; Table 2, Supplementary Table S5 and Supplementary Figure S6). Similarly, there was no evidence of colocalization of the genetic association signals between VTE and liver β2GPI (APOH) mRNA expression, or between VTE and plasma β2GPI (APOH) levels at the *APOH* locus (maximum PP4 = 0.069; Supplementary Table S5). Thus, there was no evidence that the genetic association at the *APOH* locus for anti-β2GPI was mediated by changes in β2GPI (APOH) expression, and none of the 17 non-coding variants met at least two prioritization criteria. Therefore, we did not prioritize these as causal candidates.

**Table 2.**
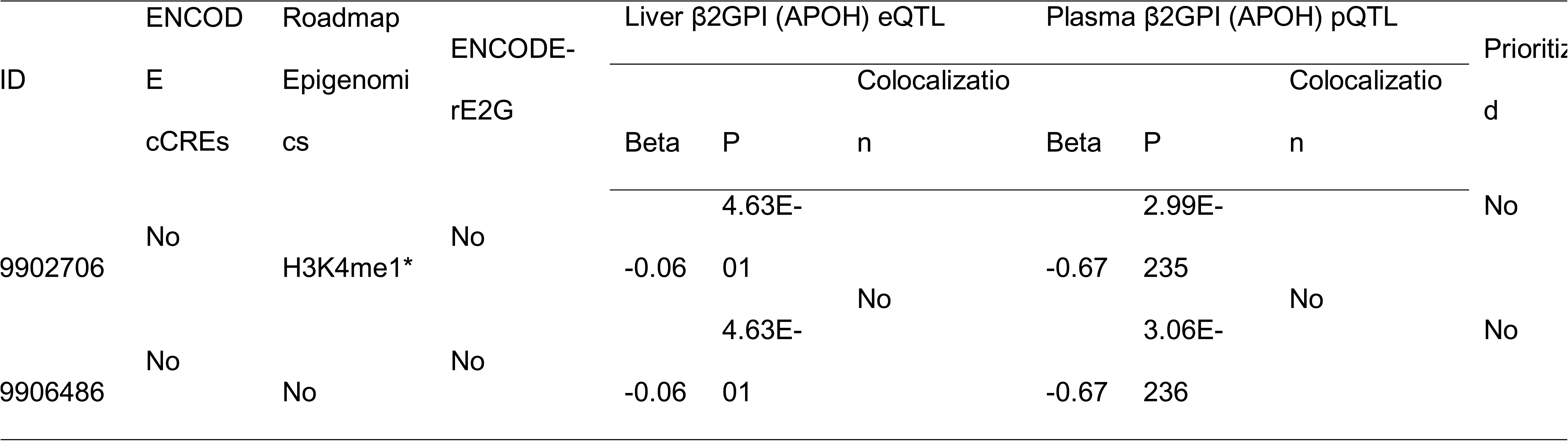
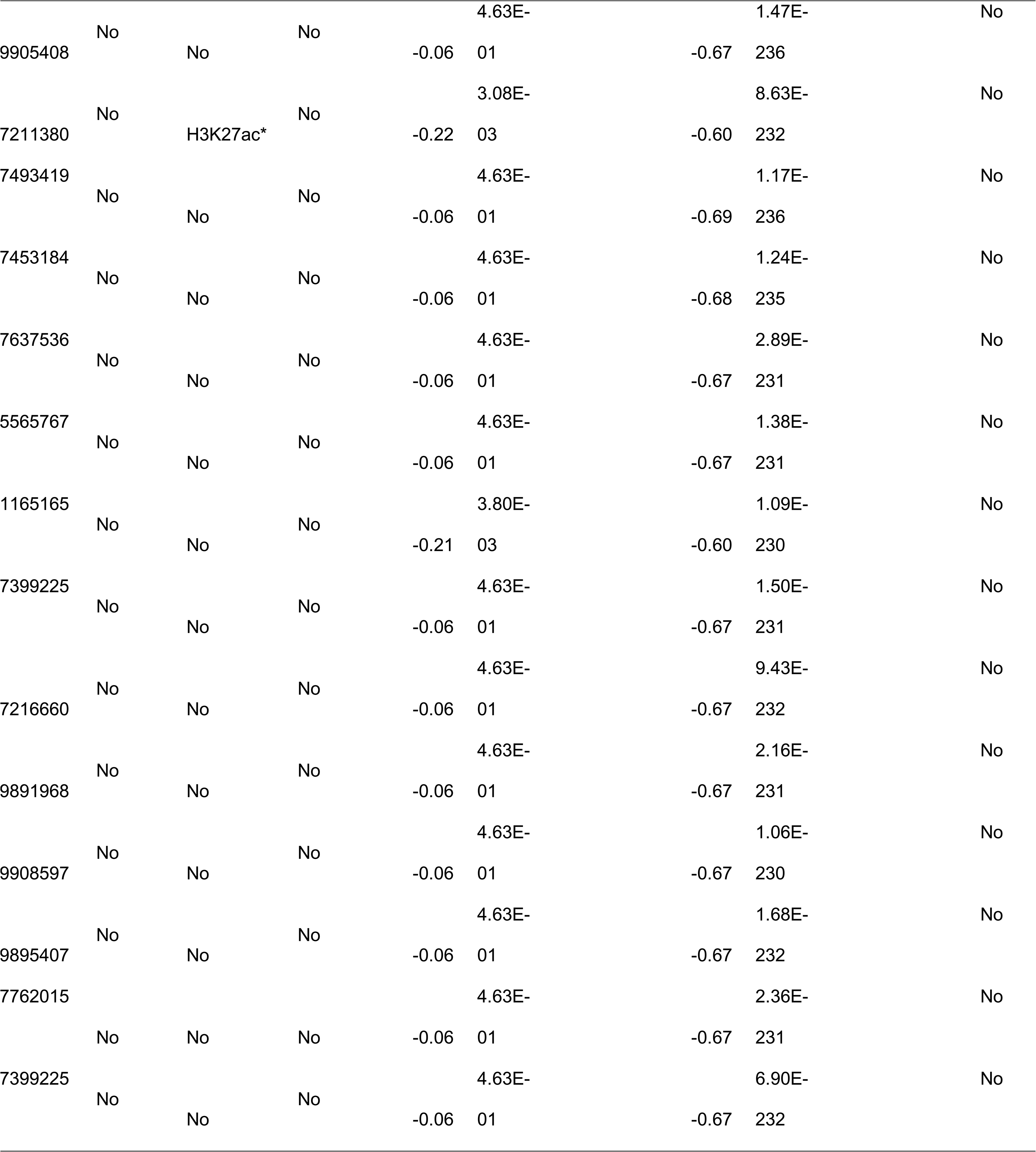
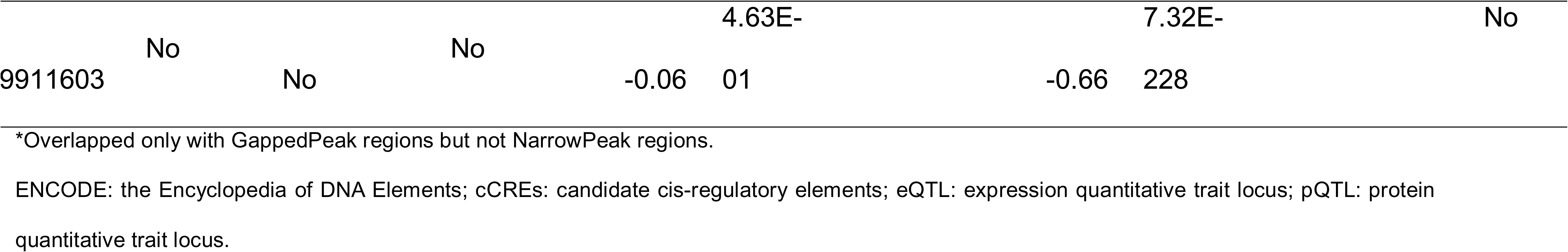
Among non-coding variants in the 95% credible sets, rs9902706 and rs7211380 overlapped with the epigenetic marks H3K4me1 and H3K27ac, respectively. None of the variants overlapped ENCODE candidate cis-regulatory elements (cCREs) or predicted to regulate β2GPI (APOH) expression in liver tissue or cell lines. All of the variants were associated with reduced β2GPI (APOH) levels in either the liver eQTL or plasma pQTL datasets, and genetic associations between anti-β2GPI antibody levels and either β2GPI (APOH) eQTL or pQTL signals did not colocalize. Overall, none of the non-coding variants were prioritized as putative mediators of the paradoxical relationship between genetically determined anti-β2GPI antibody levels and venous thromboembolism (VTE).

The variant rs1801690 is a missense variant (W335S) in the *APOH* gene. We used nine prediction tools to evaluate the functional impact of W335S: SIFT^17^, MutationAssessor^18^, PROVEAN^19^, CADD^20^, DANN^21^, LIST-S2^22^, ESM-1b^23^, AlphaMissense^24^, and PrimateAI-3D^25^. Because W335S is a common variant and not expected to cause Mendelian diseases, we specifically selected prediction tools that were not trained on datasets containing Mendelian disease labels. Among the nine variant impact prediction tools, seven (78%) inferred a deleterious effect, including SIFT, PROVEAN, CADD, DANN, ESM-1b, MutationAssessor (medium probability or deleterious), and AlphaMissense (pathogenic). The remaining two prediction tools (22%), LIST-S2 and PrimateAI-3D, classified the variant as tolerated or benign (Supplementary Table S6). These results suggest that, despite being relatively common in the general population, W335S likely meaningfully alters the structure and function of β2GPI (APOH).

Taken together, only the missense variant rs1801690 (W335S) was further prioritized as a candidate causal variant.

### Molecular dynamics

We performed molecular dynamics for both the WT and the W335S variant of β2GPI, starting from both our theorized circular model and from the crystal structure 1C1Z.^11^ We then combined all of the trajectories and performed a PCA and clustering analysis on this new single trajectory. A K-Means clustering suggested that using six clusters captured the majority of the distinct conformational states (Supplementary Figure S7). By measuring the root mean square deviation (RMSD) with respect to the cluster mid-points (Supplementary Figure S7), we confirmed that the structures placed into each cluster by the algorithm were highly similar to each other with an average RMSD of ∼ 10 Å. The radius of gyration (R_G_) of each cluster for each model was also highly similar, further confirming the structural similarity within each cluster for each model. Finally, measuring the RMSD of each cluster to the WT circular starting point or the WT linear starting point, respectively, showed that the linear cluster obtained from the circular starting points was highly similar to the linear structure obtained from the linear starting points. Cluster 1 was identified as the starting O-Shape; clusters 2 and 4 represented two distinct transition states towards cluster 3 (J-Shape 1), which then lead to either cluster 5 (J-Shape 2) or cluster 6 (S-Shape). In both the WT and the W335S variants, β2GPI gradually moved away from the starting O-Shape and extended into several distinct linear shapes (Figure 4), including one conformation highly similar to the established J-Shape. The simulations started from the 1C1Z structure remained in this cluster throughout their entire simulated time. The gradual shift from O-Shape to linear shapes occurred at a slightly slower speed with the W335S variant (Supplementary Figure S8), but the final structures were similar at the full protein level, except for one S-Shape cluster observed only in the WT simulations. The main difference among the 3 linear shapes (J-Shape 1, J-Shape 2 and S-Shape) lay within the DII to DIII interface at which the protein twisted in different directions. We then extracted residue level properties for each of the clusters and focused on previously identified regions of interest.

**Figure 4.**
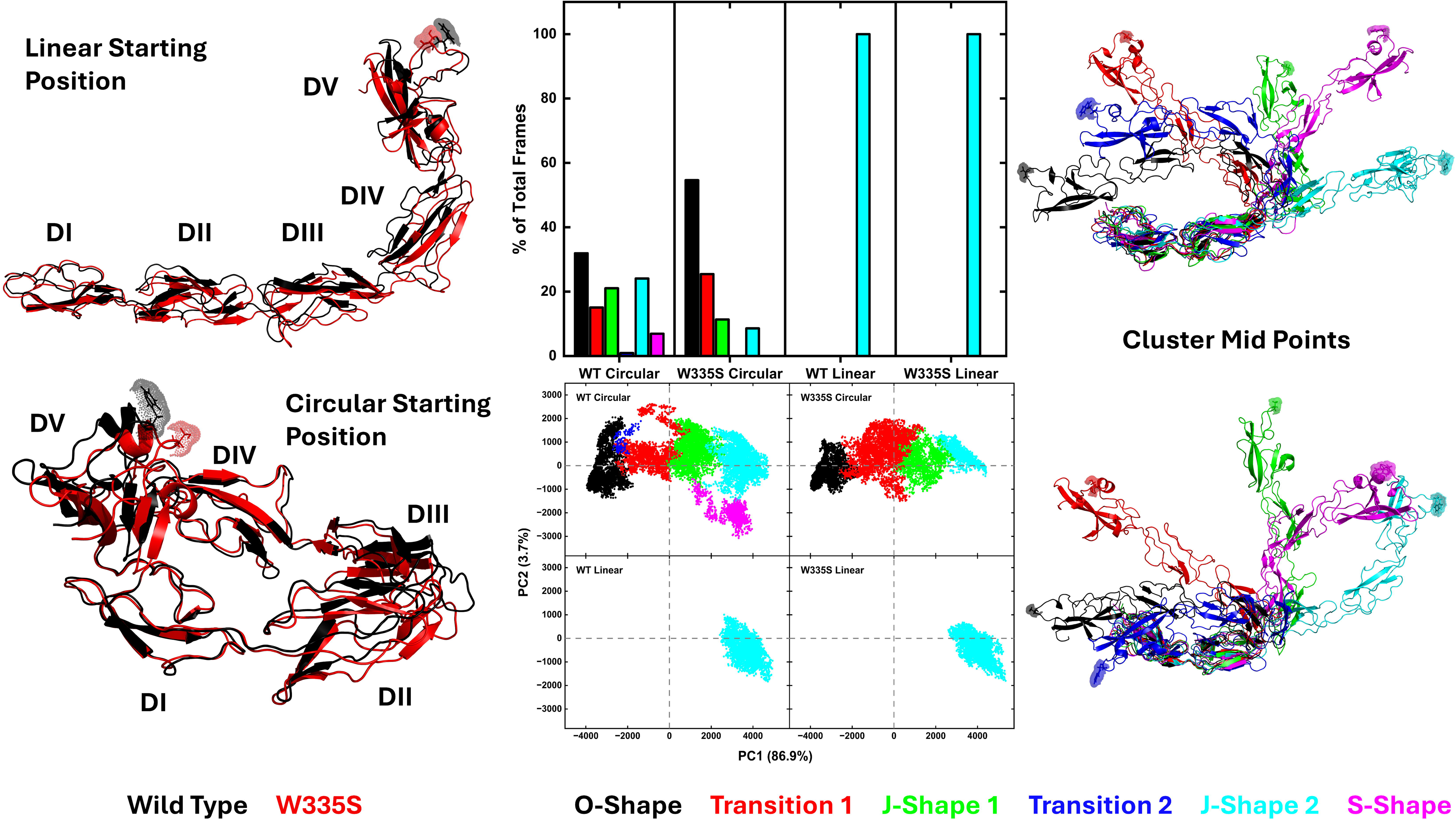
<u>Left:</u> Cartoon representation of the starting positions for the molecular simulations of this study. <u>Middle:</u> Cluster distribution after PCA and clustering analysis, with PC1 vs PC2 plots for each model. <u>Right:</u> Cluster mid-points, i.e. representative structure from each cluster.

To evaluate whether our simulations reflected the known properties of W335S^26^, we analyzed DV regions responsible for phospholipid binding. We focused on residues previously identified as key for phospholipid interaction: K269-K270^27^ and three regions described by Kolyada et al.^28^: aPL loop 1 (K303-K305-K306), aPL loop 2 (L332-F334-W335), anti-A1 loop (K301-K327-K336). It is worth noting that aPL loop 2 includes the site of the missense variant, W335S. Solvent accessible surface area (SASA) data were obtained for the frames within each cluster per model to identify residue-level differences between models within the same protein-level conformation. SASA data from the regions of interest within DV, including aPL loops 1 & 2, K269-K270 and anti-A1 loop, revealed that the variant had a large impact on their exposure (Figure 5). Within J-Shape 1 of W335S, both aPL loop 2 and residue 335 became more buried compared to the O-Shape (– ∼10% exposure), with K269-K270 also becoming more buried. When comparing the data from W335S relative to the WT in J-Shape 1, the aPL loop 2 and S335 were more buried (– ∼20% and – ∼50% exposure, respectively). In J-Shape 2 of W335S, aPL loop 2 became more exposed relative to the O-Shape, whereas in the WT this same loop became more buried relative to the O-Shape. Despite this, direct comparison of J-Shape 2 between W335S and WT showed that aPL loop 2 remained more buried overall in W335S, particularly at residue S335 (– ∼10% & – ∼50% exposure, respectively). And the average SASA of aPL 2 in both J-Shape 1 andJ-Shape 2 was lower in W335S than even the WT O-Shape (mean SASA 369.5 vs 436.9 vs 491.2, respectively). Finally, in the O-Shape, both aPL loops and residue 335 were more buried in W335S relative to the WT (– ∼5%, – ∼17% and – ∼47% exposure, respectively) (Supplementary Figure S9). These results demonstrate that across all three common protein-level conformations, the W335S variant has a decreased accessibility of specific phospholipid binding loops. The loop containing the variant itself became significantly more buried, with exposure reduced by up to 50%. Consistent with prior experimental observations^26^, our molecular dynamics simulations recapitulated the loss of phospholipid binding capacity in DV carrying the W335S variant.

**Figure 5.**
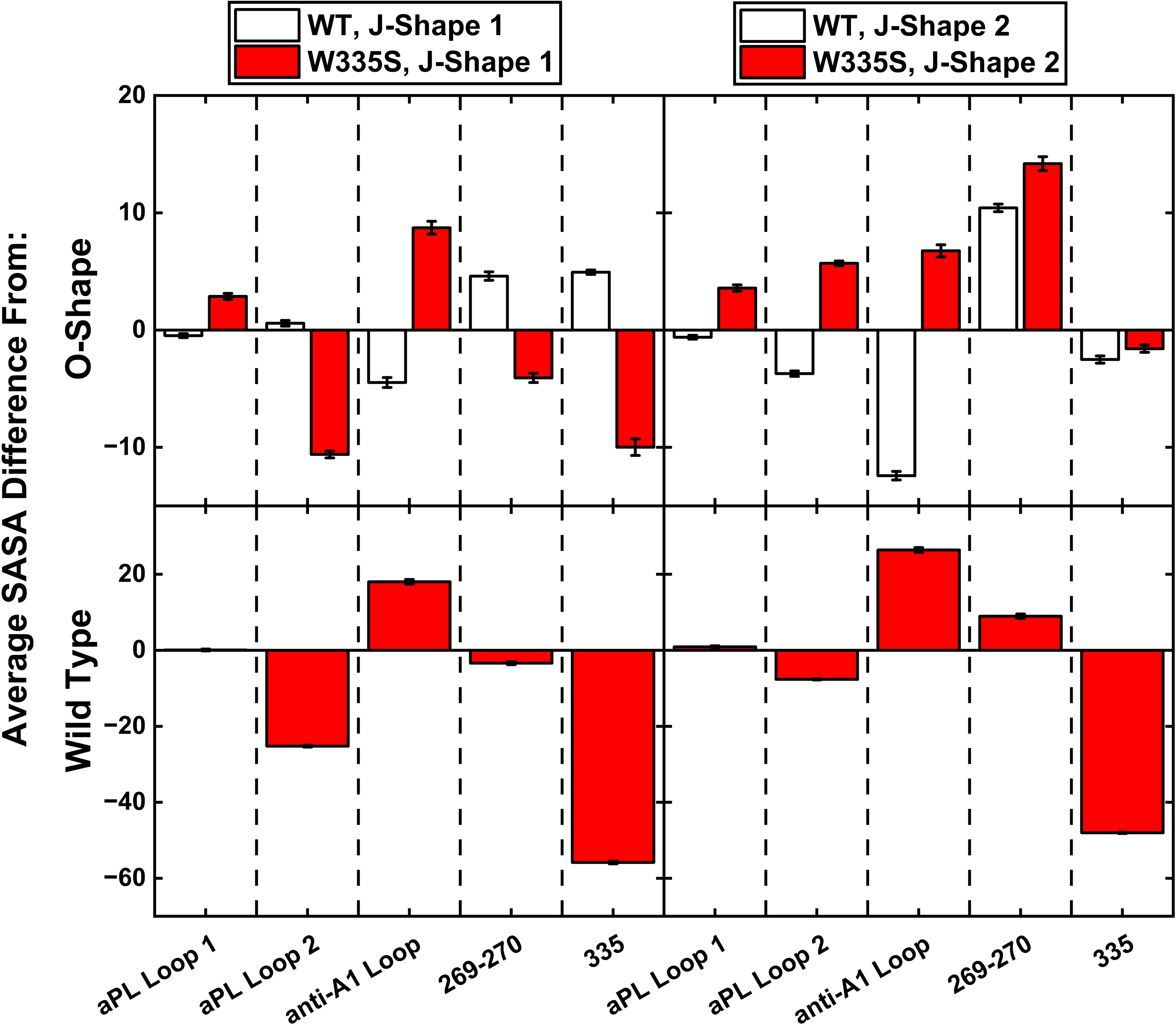
SASA data of DV areas of interest for the J-Shape 1 and J-Shape 2 clusters, relative to the O-Shape of each model (top) and relative to the same shape in the WT (bottom).

We next examined how the W335S variant alters the exposure of the established anti-β2GPI epitopes in DI and DII. We extracted residue level properties for each of the clusters and focused the analysis on the five motifs within DI & DII identified by De Moelroose et al^9^ as the main sites of autoantibody binding in APS. The results showed that consistent with the circular form hypothesis, in both the WT and variant models, the DI-DII motifs had an overall larger surface area in the linear clusters (J-Shape 1 and J-Shape 2) compared to the O-Shape (Figure 6). Specifically, motifs 2 and 4 showed an increase in SASA relative to the O-Shape (+ ∼9%, + ∼7%, respectively) for both the WT and W335S, while W335S also displayed an increase for motifs 3 & 5 (+ ∼6%, + ∼2%, respectively). When comparing the data for the same shape, W335S had larger SASA than the WT for both motifs 3 & 5 (+ ∼5% in J-Shape 1, + ∼7% / + ∼3% in J-shape 2). Of particular interest, even within the O-Shape, W335S displayed increased exposure of motifs 3 & 5, which carry the strongest antibody-reactive residues (phenylalanine and cysteine). Motif 3 includes the well-characterized 58-62 peptide, whereas motif 5 includes the DI-II linker peptide identified by Ioannou et al^8^. These findings suggest that W335S may enhance the accessibility of pathogenic epitopes, providing a plausible mechanism for increased anti-β2GPI antibody production.

**Figure 6.**
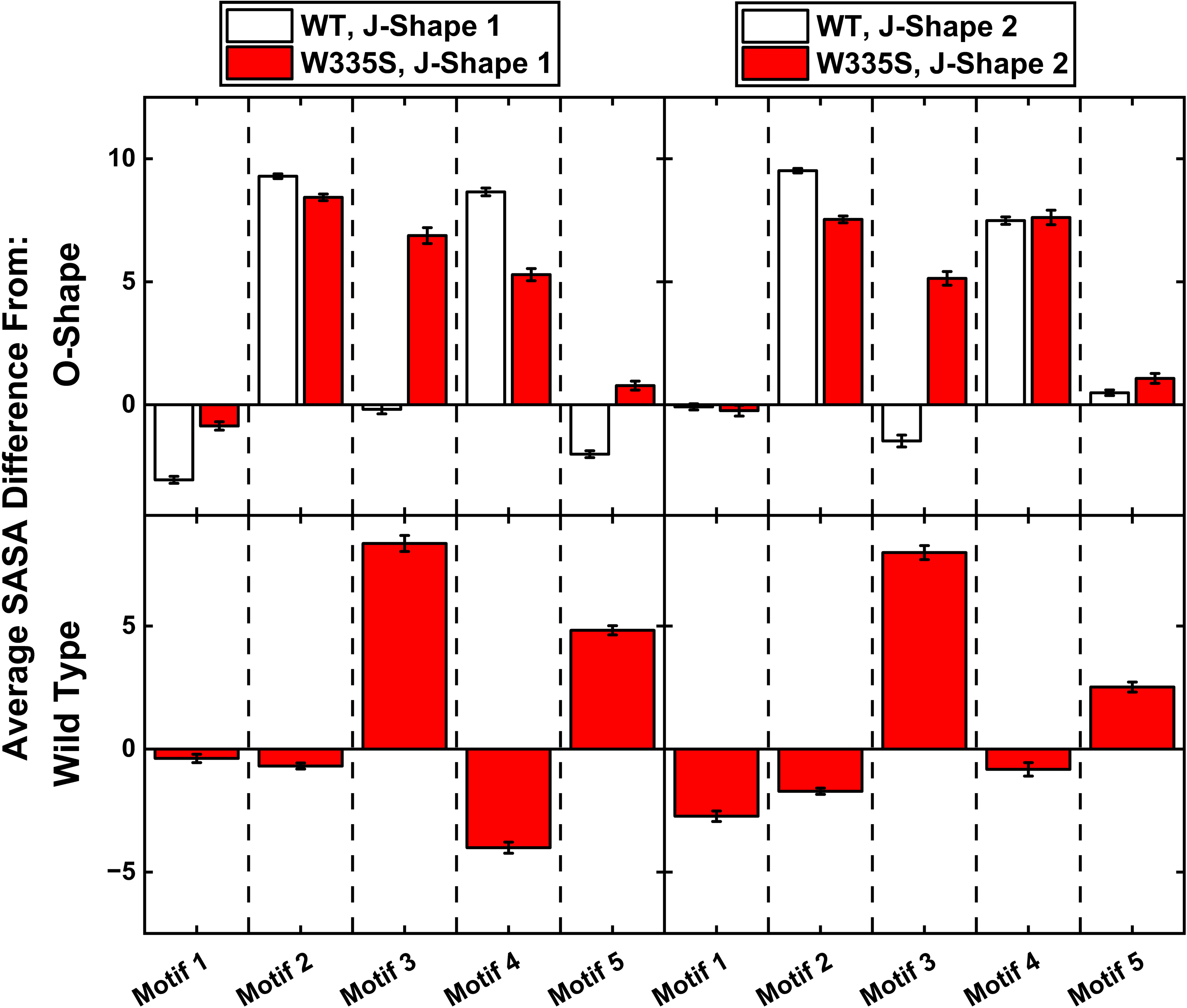
SASA data of DI-II areas of interest for the J-Shape 1 and J-Shape 2 clusters, relative to the O-Shape of each model (top) and relative to the same shape in the WT (bottom).

### Structure-based prediction with DiscoTope

We next corroborated the SASA-based observations using structure-based epitope prediction with DiscoTope 3.0. We extracted 100 frames randomly from the O-Shape, J-Shape 1 and J-Shape 2 clusters from both the WT and the W335S models. These frames were analyzed using DiscoTope 3.0 and the data averaged for each residue over the 100 frames. The results recapitulated the predicted antigenicity of almost all five motifs, especially motifs 2, 3 & 4 which were more exposed in the linear clusters relative to the O-Shape (Figure 7). The increased antigenicity was further amplified for the W335S variant. Furthermore, within each conformation, including the O-Shape, the W335S model showed higher predicted antigenicity than the WT, with the most notable increase observed in motif 3, which encompasses the established R59-R62 epitope.

**Figure 7.**
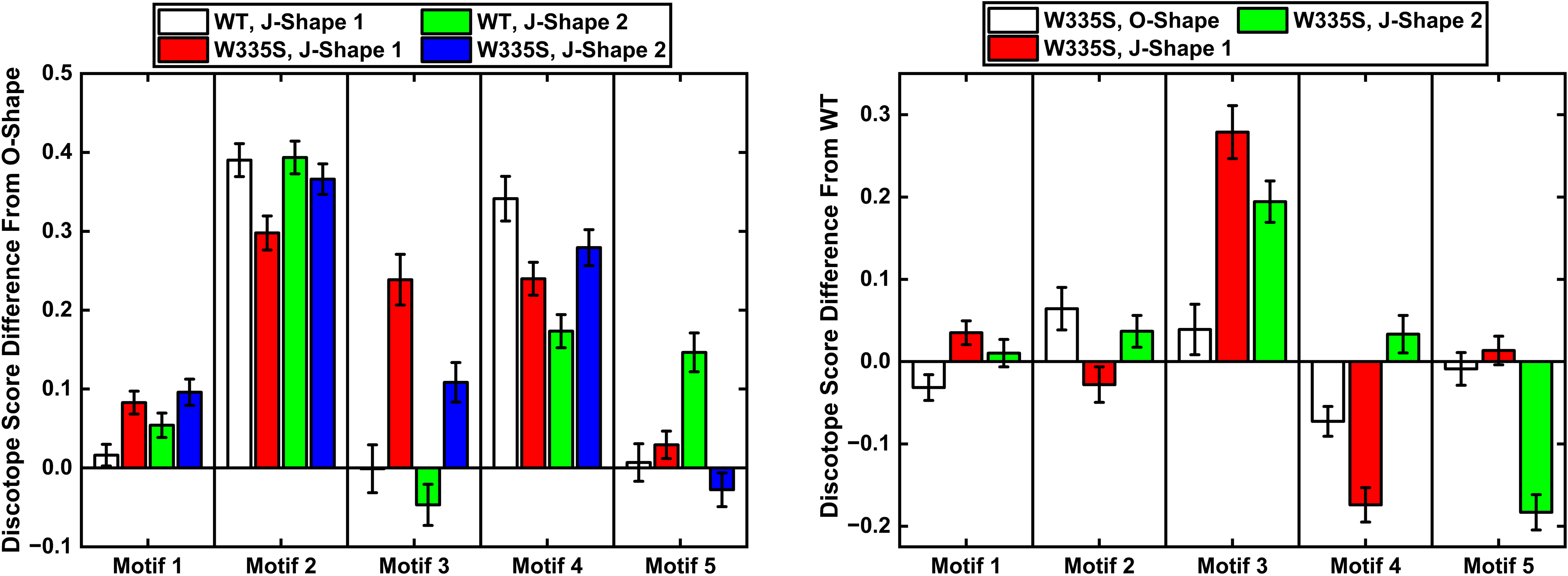
Average DiscoTope scores of the antibody binding motifs relative to the O-Shape (left), and relative to the same shape in the WT (right).

## Discussion

We conducted a multi-ancestry GWAS of serum total anti-β2GPI antibody levels in 5,969 individuals and replicated the previously reported association at the *APOH* locus.^14^ Unexpectedly, genetically determined increases in anti-β2GPI levels driven by *APOH* locus polymorphisms were associated with a reduced risk of VTE. Through colocalization, statistical fine-mapping and functional genomics analyses, we identified a missense variant, rs1801690 (W335S), as the only plausible causal variant within the credible set mediating this paradoxical relationship. Integrating prior experimental evidence, molecular dynamics simulations, and structure-based antigenicity predictions, we propose that W335S enhances autoantibody epitope accessibility in DI and DII while impairing phospholipid binding in DV, thereby promoting autoantibody formation and reducing thrombotic risk.

Given the well-established prothrombotic role of anti-β2GPI antibodies, we undertook multiple analyses to confirm the robustness of our unexpected findings. Our primary GWAS was conducted using total anti-β2GPI antibody levels as a quantitative trait, and the association was independently replicated in both positive anti-β2GPI IgM and IgG, which were measured separately from total antibody levels. To further validate our results, we replicated the Mendelian randomization findings using the lead variant previously reported in a GWAS of anti-β2GPI IgG positivity conducted in 4,163 individuals of German descent.^14^ Finally, we performed colocalization analyses to confirm that the genetic associations of anti-β2GPI antibody levels and VTE in the *APOH* locus were driven by a shared causal genomic signal.

MR typically assumes that the instrument influences the outcome solely through the exposure.^29^ Given the established thrombogenic role of anti-β2GPI, our paradoxical finding that genetic variants increasing antibody levels decrease VTE risk suggests a violation of this assumption (horizontal pleiotropy). Specifically, the causal variant affects VTE risk through a pathway independent of anti-β2GPI antibodies. Because the cis-regulated product of *APOH* is β2GPI, the autoantigen of anti-β2GPI, we initially hypothesized that the causal variant increased β2GPI expression, as higher autoantigen levels can promote autoantibody production, while β2GPI is a known anticoagulant that could directly reduce VTE risk.^2,30,31^ However, our data did not support this hypothesis: the non-coding variants in the 95% credible sets were associated with reduced, rather than increased, plasma β2GPI levels. We further demonstrated genetic association signals for anti-β2GPI antibody levels and β2GPI levels, either as liver mRNA expression or plasma protein abundance, did not colocalize. Furthermore, the regulatory role of these non-coding variants was also not supported by ENCODE-rE2G predictions.^32^ Taken together, these findings indicate that the causal genomic signal underlying anti-β2GPI antibody levels is unlikely to be responsible for changes in β2GPI expression. Therefore, the GWAS lead variant, rs1801690 (W335S), emerged as the sole remaining candidate causal variant. Notably, despite being a common missense variant, seven of nine variant effect predictors classified it as deleterious. This includes AlphaMissense, which was explicitly fine-tuned on population allele frequencies and is therefore “biased” toward predicting benign for common variants.^24^ Its deleterious prediction for W335S thus supports the likelihood that this substitution has a substantial impact on β2GPI structure and function.

An association between W335S and lower plasma β2GPI levels has been previously described.^33,34^ However, these studies lacked the sample size and locus-wide dense variant coverage necessary to determine whether the relationship was causal. By leveraging modern eQTL and pQTL resources, as well as Bayesian colocalization framework, we found no evidence that reduced β2GPI levels are driven by W335S itself, but instead by a distinct genomic signal in LD with W335S.

W335S (also reported as W316S in studies that exclude the signal peptide from residue numbering) is located within the aPL loop 2 of DV, a critical region through which β2GPI inserts into anionic phospholipid membranes. Substitution of the hydrophobic tryptophan with a hydrophilic serine at this position has been experimentally shown to abolish the ability of β2GPI to bind phospholipids.^26^ This loss of phospholipid-binding capacity provides a biologically plausible explanation for our finding that the genomic signal at the *APOH* locus is associated with reduced VTE risk. Consistent with these experimental data, our molecular dynamics simulations recapitulated the loss of phospholipid-binding capacity at residue-level resolution.

Genetically driven increased antigen levels leading to higher autoantibody production has been reported in other autoimmune conditions, such as PR3-associated vasculitis and PLA2R-associated membrane nephropathy.^30,31^ Motivated by these observations, we proposed two hypotheses to explain how W335S increases anti-β2GPI antibody production. The first hypothesis was that W335S reduces the proportion of the circular (O-Shape) conformation of β2GPI in circulation, thereby shifting the equilibrium toward more linear (J-Shape) conformations and increasing epitope exposure. The second hypothesis was that W335S, located in DV, induces a global conformational change that increases antigen exposure in DI and DII, analogous to the increased antigenicity observed after removal of amino acids 318–326 from plasmin cleavage.^10^ It is important to note that the existence of the circular conformation remains debated, with supportive evidence from molecular dynamics simulations but conflicting experimental data.^11–13^ To evaluate both hypotheses, we performed molecular dynamics simulations initiated from both the theorized O-Shape and the crystallographic J-Shape. The results showed that W335S did not reduce the representation of the circular conformation, arguing against our first hypothesis. Instead, our findings supported the second hypothesis: across all conformations, W335S consistently increased SASA for motifs 3 and 5, which contain the most strongly antibody-reactive residues. Complementary structure-based epitope prediction using DiscoTope 3.0 also demonstrated greater antigenicity in these motifs for W335S compared to the wild type.^35^ Taken together, these *in-silico* analyses provide a plausible mechanistic explanation for the elevated anti-β2GPI antibody levels associated with W335S in our genetic association study.

Our study has potentially important clinical implications. Anti-β2GPI testing is routinely performed in the evaluation of patients with recurrent pregnancy loss and those with systemic autoimmune rheumatic diseases such as systemic lupus erythematosus (SLE), including in individuals without a history of thrombosis.^36,37^ The observation that the W335S variant is associated with higher anti-β2GPI antibody levels but lower risk of VTE suggests that the presence of anti-β2GPI antibodies may not uniformly confer increased thrombotic risk across all genetic backgrounds. This finding may have implications for thrombotic risk stratification and clinical management in individuals who are anti-β2GPI-positive and carriers of W335S. It is also important to note that our analyses were conducted in a general population cohort rather than in populations at high risk for autoimmune, thrombotic and/or obstetric events. A recent GWAS of APS identified two genome-wide significant and several suggestive loci; however, none were located within *APOH*, albeit in a relatively small sample of 482 cases. These findings suggest that, although anti-β2GPI antibodies are central to APS, additional immunologic pathways contribute to clinical disease.^38^ The role of W335S in individuals with a higher baseline propensity for autoantibody production or thrombosis, such as patients with SLE or other systemic autoimmune rheumatic diseases, warrants further investigation.

Our study has several limitations. First, measurements of total anti-β2GPI, anti-β2GPI IgM, and anti-β2GPI IgG in MESA were not originally designed for genetic association studies. Quantitative values were available for all participants only for total anti-β2GPI, whereas IgM and IgG isotypes were assayed only in individuals in the top 20th percentile of total anti-β2GPI. As a result, statistical power for the dichotomized IgM and IgG traits was limited, and they were performed as secondary analyses. Nevertheless, the main genetic association and MR findings were replicated in anti-β2GPI IgM and IgG. Second, although statistical fine-mapping identified rs1801690 (W335S) as the variant with the highest posterior inclusion probability (PIP = 0.49), this probability was not definitive. The final prioritization of rs1801690 therefore relied on integrating fine-mapping with functional genomic evidence from eQTL, pQTL, and epigenomic predictions. We cannot definitively exclude the possibility that the paradoxical effect is mediated by an unmeasured variant, although this is less likely given the dense imputation-based coverage across the locus. Third, the *APOH* locus showed only suggestive association with VTE. However, genome-wide statistical significance of the instrument in the outcome dataset is not required for a valid causal inference through MR.^29^ Likewise, genome-wide statistical significance is not required to infer a shared causal variant in Bayesian colocalization analyses, as inference in this framework is based on posterior probabilities rather than null-hypothesis significance tests.^39^ Fourth, our mechanistic proposal is based on molecular dynamics simulations and structure-based epitope prediction but without experimental validation, and thus should be interpreted as hypothesis-generating. Finally, we did not measure anti-DI antibodies, so it is possible that the increase in total anti-β2GPI may reflect autoantibodies targeting other domains.

In conclusion, we discovered that polymorphisms at the *APOH* locus associated with increased anti-β2GPI antibody levels paradoxically protect against VTE. We prioritized a missense variant, rs1801690 (W335S), as the most likely causal variant mediating this effect. Integrating molecular dynamics simulations, structure-based epitope prediction, and prior experimental evidence, we propose a mechanistic model in which W335S abolishes phospholipid binding in DV, thereby reducing thrombotic risk, while inducing global conformational changes that increase epitope exposure in DI and DII and enhance autoantibody production. These findings raise the possibility that thrombotic risk may vary among anti-β2GPI-positive individuals depending on their *APOH* genotype, highlighting the need for future research on genotype-guided risk stratification.

## Methods

### Study population

The Multi-Ethnic Study of Atherosclerosis (MESA) is a prospective cohort study of 6,814 community-dwelling adults aged 45-84 years from four racial/ethnic groups (White, African American, Hispanic, Chinese American) recruited from six U.S. sites (Forsyth County, North Carolina; northern Manhattan and the Bronx, New York; Baltimore City and Baltimore County, Maryland; St. Paul, Minnesota; Chicago, Illinois; and Los Angeles, California) between 2000-2002.^40^ The cohort is racially and ethnically diverse, comprising approximately 38% White, 28% African American, 22% Hispanic, and 12% Chinese American participants.

### Phenotype measurement

Anti-β2-glycoprotein I (anti-β2GPI) antibodies were measured by enzyme-linked immunosorbent assay (ELISA) using commercial kits (TheraTest Labs Inc, Lombard, IL, USA), as previously described.^41^ The assays are FDA-approved for clinical use and performed in two stages: an initial screening test followed by isotype-specific testing for positive results. The screening assay is a solid-phase enzyme immunoassay that detects total anti-β2GPI antibodies (IgG, IgM, and IgA combined). Plasma samples with values exceeding 10.5 units underwent additional testing to quantify individual isotypes (IgG, IgM, and IgA).

### Genotyping, quality control and imputation

The MESA participants were genotyped using the Affymetrix Human SNP array 6.0.^42^ Quality control (QC) procedures included filters of per-variant genotyping rate ≥ 95%, per-individual genotyping rate ≥ 98%, and minor allele frequency (MAF) ≥ 0.01. Sex of each participant was inferred from sex chromosome markers, and individuals with discrepancies between imputed and recorded sex were excluded.

Principal component analysis (PCA) was first performed on linkage disequilibrium (LD)–pruned overlapping variants in unrelated individuals from the 1000 Genomes Project.^43^ The resulting PCs defined the reference space into which PCs from MESA participants were subsequently projected. Ancestry inference was then conducted using a random forest classifier trained on the top 10 PCs of the 1000 Genomes reference individuals. Within each inferred ancestry group, a second round of PCA was carried out using SmartPCA to refine population structure.^44^ Participants who were identified as PC outliers within their ancestry group were excluded from further analyses.

Genotype imputation was performed using the NHLBI Trans-Omics for Precision Medicine (TOPMed) reference panel (Freeze 10) on the TOPMed Imputation Server.^45,46^ Imputation was carried out using the Eagle v2.4 algorithm for pre-phasing and Minimac4 for imputation, with default server parameters.^47,48^ A total of 17,354,364 high-quality common variants (R^2^ ≥ 0.8 and MAF ≥ 0.01 in at least an ancestral group) were retained for downstream GWAS analyses.

### Genome-wide association analysis

The primary outcome was total anti-β2GPI antibody levels, analyzed as a quantitative trait. Antibody values were inverse normal transformed (INT) prior to genetic association testing. Genome-wide association studies (GWAS) were conducted within each ancestry group using REGENIE v4.1, which implements a two-step ridge regression framework.^49^ In Step 1, whole-genome regression on high-quality genotyped variants was used to generate polygenic predictions under a leave-one-chromosome-out (LOCO) scheme to account for relatedness and population structure. In Step 2, association testing of imputed variants was performed using linear regression with LOCO predictions included as an offset. Covariates were age, sex, study site, smoking status, educational level, and the top 10 ancestry-specific principal components. Ancestry-specific GWAS results were then combined via fixed-effect inverse-variance weighted meta-analysis using METAL.^50^

We performed secondary GWAS analyses for anti-β2GPI antibody isotypes (IgM, IgG, and IgA). Since isotypes were only measured in individuals with total anti-β2GPI antibody levels in the top 20th percentile, treating them as quantitative traits was not feasible. Instead, each isotype was evaluated as a binary trait using multiple positivity thresholds (90th, 95th, 98th, and 99th percentiles, as well as the manufacturer’s cutoff), with GWAS performed using REGENIE as described above. Given that the LOCO scheme may lead to test statistic deflation in small sample sizes, logistic regression was conducted within each ancestry group restricted to unrelated individuals, without LOCO.

### Statistical fine-mapping

To identify candidate causal variants at the *APOH* locus associated with total anti-β2GPI antibody levels, we performed statistical fine-mapping and generated 95% credible sets. Our primary approach used the Sum of Single Effects (SuSiE) regression, implemented in the R package susieR, applied to multi-ancestry meta-analysis summary statistics.^51^ As a complementary analysis, we also applied MESuSiE, a multi-ancestry fine-mapping method based on the SuSiE framework that jointly models both shared and ancestry-specific causal variants.^52^ Although MESuSiE incorporates ancestry-specific information, the modest sample sizes within individual ancestry groups yielded weaker association signals, limiting fine-mapping resolution. For both SuSiE and MESuSiE, in-sample LD from unrelated individuals was used as the reference panel.

MESuSiE was restricted to variants that were present in all ancestral groups. Because the meta-analysis lead variant (rs1801690) had a MAF of 0.0099 in AFR, fine-mapping with MESuSiE was performed using ancestry-specific variants with MAF ≥ 0.009.

### Mendelian randomization and colocalization

Anti-β2GPI antibody is an established cause of venous thromboembolism (VTE) through antiphospholipid syndrome (APS). To evaluate the relationship between genetically determined anti-β2GPI antibody levels at the *APOH* locus and VTE risk, we obtained summary statistics from a recently published GWAS meta-analysis of VTE, which included 81,190 cases and 11,419,671 controls of European ancestry from six cohorts.^15^ We then performed two-sample Mendelian randomization (MR) using rs1801690 as the instrumental variable (IV), with total anti-β2GPI antibody levels as the exposure and VTE as the outcome. Causal estimates were obtained using the Wald ratio method under the NOME (No Measurement Error) assumption, as implemented in the TwoSampleMR R package (version 0.6.6). To assess whether the causal relationship detected by MR was attributable to shared causal variants, we performed colocalization analysis using the coloc.susie function from the coloc R package (version 5.2.3).^53^ LD was estimated using in-sample genotypes for the anti-β2GPI GWAS and 1000 Genomes Europeans for the VTE GWAS.^43^ We considered evidence for a shared causal variant if the posterior probability for colocalization (PP4) exceeded 0.9, and values between 0.5 and 0.9 were considered suggestive.

### Causal variant prioritization

Because the paradoxical association of genetically determined anti-β2GPI antibody levels with reduced VTE risk likely reflects a violation of standard MR assumptions, we hypothesized that this relationship is mediated not by the antibodies themselves but by a downstream consequence of the causal variant in the *APOH* locus—most plausibly β2GPI (APOH), the autoantigen of anti-β2GPI antibodies.

To identify the causal variants within the 95% credible sets that are biologically plausible mediators of the paradoxical association between higher anti-β2GPI antibody levels and lower VTE risk, we focused on (1) non-coding variants predicted to increase β2GPI (APOH) expression, and (2) non-synonymous protein-coding variants. Increased β2GPI (APOH) expression is a plausible mechanism because β2GPI (APOH) is an established anticoagulant, and genetically determined higher autoantigen levels have been reported to drive autoantibody production in certain contexts, such as proteinase 3 (PR3)^30^ and phospholipase A2 receptor (PLA2R)^31^. Evidence supporting causality through increased APOH expression was assessed using four criteria, evaluated in relevant tissues (liver for mRNA expression and epigenetic studies, plasma for protein levels) where applicable: (1) overlap of the variant with regulatory regions inferred from the Encyclopedia of DNA Elements (ENCODE)^54^ or Roadmap Epigenomics Project^55^; (2) overlap with regulatory regions predicted by ENCODE-rE2G to regulate β2GPI (APOH) expression;^32^ (3) association of the variant with increased β2GPI (APOH) mRNA expression or protein levels; and (4) evidence of colocalization between genetic association signals for anti-β2GPI antibody levels and β2GPI (APOH) mRNA expression or protein levels. Non-coding variants meeting at least two of these criteria were further prioritized as candidate causal variants. Summary statistics for the β2GPI (APOH) liver expression quantitative trait locus (eQTL) were obtained from the Genotype-Tissue Expression (GTEx, version 8)^56^, and for the β2GPI (APOH) plasma protein quantitative trait locus (pQTL) from the UK Biobank^57^. Detailed descriptions of how each criterion was evaluated are provided in the Supplementary Note. To evaluate the functional impact of protein-coding variants, we applied nine prediction tools: SIFT^17^, MutationAssessor^18^, PROVEAN^19^, CADD^20^, DANN^21^, LIST-S2^22^, ESM-1b^23^, AlphaMissense^24^, and PrimateAI-3D^25^. Because our candidate variants from GWAS were common and not expected to cause Mendelian diseases, we specifically selected prediction tools that were not trained on datasets containing Mendelian disease labels, such as ClinVar^58^ or Human Gene Mutation Database (HGMD)^59^. Detailed descriptions of the selected prediction tools are provided in the Supplementary Note.

### Sensitivity analysis

GWAS was initially restricted to variants with ancestry-specific MAF ≥ 0.01. However, the lead variant at the APOH locus (rs1801690), identified in the multi-ancestry meta-analysis under this threshold, had a MAF of 0.0099 in AFR. Therefore, sensitivity analyses at this locus were performed by including ancestry-specific variants with MAF ≥ 0.009 in both GWAS and downstream post-GWAS analyses (Mendelian randomization, colocalization, and statistical fine-mapping).

We also performed colocalization analyses between positive anti-β2GPI IgM (90th percentile cutoff and manufacturer cutoff) and IgG (90th percentile cutoff) from our MESA study and VTE. In addition, to extend the analysis to an external GWAS, the *APOH* locus was previously reported as genome-wide significant for positive anti-β2GPI IgG (99th percentile cutoff) in a GWAS of 4,163 individuals of German descent.^14^ We performed two-sample MR using positive anti-β2GPI IgG as the exposure, the lead variant in this dataset (rs8178848) as the IV, and VTE as the outcome. Because full summary statistics from this study were not available, colocalization analysis could not be performed.

### Preparing molecular variants of **β**2GPI for molecular dynamics simulations

A theorized circular model of β2GPI was generated starting from the linear crystal structure (PDBID: 1C1Z,^3^ as described in previously.^11^ Briefly, experimental literature data suggests either a head-to-head closure between DI and DV or a closure between DV and the DI-II interface. An affinity analysis was run between DI-II and DV using molecular dynamics simulations and a total of 24 positions were generated. The system was minimized, solvated, and equilibrated; following which close contact probabilities between domains were calculated by counting the occurrences of distances below 0.24 nm between all atoms. The most specific interaction was seen between K295 and E42, and ring closure was assumed to occur at this contact point. To close the ring, a flat-bottom potential was applied between the amino group nitrogen of K295 and the carboxyl group carbon of E42. This closed ring structure was not stable after removing the restraint, but the ring could be further stabilized by the inclusion of branched glycans as described by Kondo *et al*.^60^ This theorized circular model acted as the basis for subsequent simulations. Protonatable residues were edited on CHARMM-GUI^61^ for correct ionization at pH 7.4.

The missense variant identified in the cohort was modelled based on this structure, with the tryptophan residue 335 mutated to serine (W335S) in the Charmm-GUI PDB reader module.^61^ Both variants had four biantennary sialylated glycans (Man_3_GlcNAc_2_ core and two NeuNAc.Gal.GlcNAc antennae) attached to N162, N183, N193, N253 as detected by Kondo et al.^60^ and Baerenfaenger et al.^62^ Simulations were also performed using the crystal structure 1C1Z as the starting point, both for the wild type (WT) and the W335S variant.

### Molecular dynamics

Ring opening trajectories were simulated using the nanoscale molecular dynamics 2 (NAMD2) program.^63^ The simulation conditions were set using the CHARMM-GUI: temperature 303.15 K, pH 7.4, ionic strength 0.15 M simulated with NaCl ions, pressure 1 bar, in explicit solvent with TIP3P water molecules^64^ and counter-ions to neutralize charge in the simulation box. The box was set to leave at least 1 nm distance from the protein on each axis. The CHARMM36 forcefield was used.^64–66^ The particle mesh Ewald algorithm with a spacing of 0.1 nm was applied to calculate long-range electrostatic forces (with the neighbor list updated every 40 fs). The short-range electrostatic and Van-der-Waals interactions were calculated with a cutoff of 1.2 nm. The Van-der-Waals interactions were smoothly switched off at 1 nm by a force-switching function (37). SHAKE was used to constrain all bonds involving hydrogen atoms. The timestep used was 2 fs, with coordinates saved every 100 ps. All systems, WT and W335S, circular and linear, were equilibrated in NVT.^63^ Positional restraints for carbon alpha backbone atoms were applied to ensure gradual equilibration of the system over 100,000 steps (200 ps). Simulations were run in NPT for a total of 150 ns, with the Langevin coupling coefficient set to 1 ps^-1^ and a Nose-Hoover Langevin piston was used to maintain constant pressure,^67,68^ with a piston period of 50 fs and a piston decay of 25 fs.

### PCA for molecular dynamics

Coordinate PCA was performed on a combined trajectory of the 10 repeats of WT, 10 repeats of W335S, 3 repeats of WT and 3 repeats of W335S started from the 1C1Z structure. This was achieved with the R library Bio3D ^69^ using the “average” algorithm for hierarchical clustering, using the first 4 principal components and splitting the frames into 6 clusters in the PCA dimensional space. By combining all the simulations into one single trajectory and performing PCA and clustering on this combined trajectory, we ensure the principal components used for the clustering are the same for every repeat and model and represent the composite PC weighting that best represents all the motions identified in the full dataset. This can then provide information about the distribution of the structural clusters as a function of the model and condition used in each simulation.^11,70–73^

### Root mean square deviation (RMSD), radius of gyration (R_G_), and solvent-accessible surface area (SASA) measurements

RMSD, R_G_ and SASA were measured using the visual molecular dynamics (VMD) software^74^ and tool command language (TCL) scripts to iterate over the frames and residues. RMSD was measured for the full protein, after aligning the full protein, either from the WT circular starting point, the WT linear starting point, or the mid-point of each cluster identified. RMSD is used to provide information on structural similarities between two frames, with low RMSD values indicating similar conformations and high RMSD values indicating dissimilar structures, though the same high-value RMSD can be due to two different structures owing to the averaging over the whole structure.^75^ It is therefore important to supplement this information with PCA and clustering to identify correct conformational groups. SASA was measured for every residue over the trajectories and was used to identify residues which are exposed to the solvent. This can aid in identifying potential epitope regions. This can be measured for the whole protein and for each residue individually, giving insight into the behavior of specific peptides within the structure by monitoring the SASA over time.^76^

### Structure-based prediction with DiscoTope

Using the trajectories and clustering analysis described above, we isolated 100 frames selected randomly from clusters of interest using a TCL script for VMD and uploaded these to the DiscoTope 3.0 server.^35^ DiscoTope provides a score predicting the antigenicity of each residue, and we averaged this value for each residue using the 100 random frames. We then measured the antigenicity score of five previously identified motifs within DI and DII of β2GPI, and measured the difference in antigenicity of the two linear clusters relative to the circular cluster within both models as well as the difference for the same cluster between the two models.

## Ethics statement

All participants provided informed consent to participate in genetic studies. the Institutional Review Board of Columbia University approved our studies under the protocol AAAC7385 and AAAI1632.

## Acknowledgement

This research was supported by contracts 75N92025D00022, 75N92020D00001, HHSN268201500003I, N01-HC-95159, 75N92025D00026, 75N92020D00005, N01-HC-95160, 75N92020D00002, N01-HC-95161, 75N92025D00024, 75N92020D00003, N01-HC-95162, 75N92025D00027, 75N92020D00006, N01-HC-95163, 75N92025D00025, 75N92020D00004, N01-HC-95164, 75N92025D00028, 75N92020D00007, N01-HC-95165, N01-HC-95166, N01-HC-95167, N01-HC-95168 and N01-HC-95169 from the National Heart, Lung, and Blood Institute, and by grants UL1-TR-000040, UL1-TR-001079, and UL1-TR-001420 from the National Center for Advancing Translational Sciences (NCATS). The authors thank the other investigators, the staff, and the participants of the MESA study for their valuable contributions. A full list of participating MESA investigators and institutions can be found at http://www.mesa-nhlbi.org. This paper has been reviewed and approved by the MESA Publications and Presentations Committee. Funding for SHARe genotyping was provided by NHLBI Contract N02-HL-64278. Genotyping was performed at Affymetrix (Santa Clara, California, USA) and the Broad Institute of Harvard and MIT (Boston, Massachusetts, USA) using the Affymetrix Genome-Wide Human SNP Array 6.0.

Y.L.’s work was supported by the National Institute of Arthritis and Musculoskeletal and Skin Diseases (grant K23-AR-084061) and the Columbia University Research Stabilization Award. L.L.’s work was supported by the National Institute of Diabetes and Digestive and Kidney Diseases (grant K01-DK-137031).

## Data availability

The MESA SHARe genotype and phenotype data are available through dbGAP, accession number phs000209.v13.p3. GWAS summary statistics have been deposited in Figshare under a private, reviewer-only access link (https://figshare.com/s/9705f29e7a1ce3bfd647). Upon acceptance and publication of this manuscript, the full summary statistics will be made publicly available without restriction.

The VTE summary statistics are available at https://www.decode.com/summarydata. The plasma β2GPI (APOH) pQTL summary statistics from UKBB are available at https://registry.opendata.aws/ukbppp/. The liver β2GPI (APOH) eQTL summary statistics from GTEx v8 are available at https://gtexportal.org/home/. The ENCODE cCRE data and ENCODE-rE2G predictions are available at https://www.encodeproject.org/. The Roadmap Epigenomics Project data are available at https://egg2.wustl.edu/roadmap/web_portal/. The variant effect predictions with SIFT, MutationAssessor, PROVEAN, CADD, DANN, LIST-S2, ESM-1b and AlphaMissense are available at https://www.dbnsfp.org/home. The PrimateAI-3D predictions are available at https://github.com/Illumina/PrimateAI-3D.

## Code availability

The following software and packages were used for data analysis: PLINK v.2.0 (https://www.cog-genomics.org/plink/2.0/), REGENIE v.4.0 (https://rgcgithub.github.io/regenie/), METAL v.2011-03-25 (http://csg.sph.umich.edu/abecasis/Metal/download/), TwoSampleMR v.0.6.6 (https://mrcieu.github.io/TwoSampleMR/), coloc v.5.2.3 (https://chr1swallace.github.io/coloc/), susieR v.0.14.2 (https://stephenslab.github.io/susieR/), MESuSiE v.1.0 (https://github.com/borangao/MESuSiE), nanoscale molecular dynamics 2 (NAMD2, https://www.ks.uiuc.edu/Research/namd/), DiscoTope (https://services.healthtech.dtu.dk/services/DiscoTope-3.0/)

## References

1 Schreiber, K. et al. Antiphospholipid syndrome. Nat Rev Dis Primers 4, 17103 (2018). 10.1038/nrdp.2017.103

2 McDonnell, T. et al. The role of beta-2-glycoprotein I in health and disease associating structure with function: More than just APS. Blood Rev 39, 100610 (2020). 10.1016/j.blre.2019.100610

3 Schwarzenbacher, R. et al. Crystal structure of human beta2-glycoprotein I: implications for phospholipid binding and the antiphospholipid syndrome. EMBO J 18, 6228–6239 (1999). 10.1093/emboj/18.22.6228

4 Hammel, M. et al. Solution structure of human and bovine beta(2)-glycoprotein I revealed by small-angle X-ray scattering. J Mol Biol 321, 85–97 (2002). 10.1016/s0022-2836(02)00621-6

5 de Laat, B. et al. The association between circulating antibodies against domain I of beta2-glycoprotein I and thrombosis: an international multicenter study. J Thromb Haemost 7, 1767–1773 (2009). 10.1111/j.1538-7836.2009.03588.x

6 Moghbel, M. et al. Epitope of antiphospholipid antibodies retrieved from peptide microarray based on R39-R43 of beta2-glycoprotein I. Res Pract Thromb Haemost 6, e12828 (2022). 10.1002/rth2.12828

7 Iverson, G. M. et al. Use of single point mutations in domain I of beta 2-glycoprotein I to determine fine antigenic specificity of antiphospholipid autoantibodies. J Immunol 169, 7097–7103 (2002). 10.4049/jimmunol.169.12.7097

8 Ioannou, Y. et al. Binding of antiphospholipid antibodies to discontinuous epitopes on domain I of human beta(2)-glycoprotein I: mutation studies including residues R39 to R43. Arthritis Rheum 56, 280–290 (2007). 10.1002/art.22306

9 de Moerloose, P. et al. Patient-derived anti-beta2GP1 antibodies recognize a peptide motif pattern and not a specific sequence of residues. Haematologica 102, 1324–1332 (2017). 10.3324/haematol.2017.170381

10 Bradford, H. F. et al. Plasmin cleavage of beta2-glycoprotein I alters its structure and ability to bind to pathogenic antibodies. J Thromb Haemost 23, 1864–1878 (2025). 10.1016/j.jtha.2025.02.015

11 Lalaurie, C. J. et al. The J-shape of beta2GPI reveals a cryptic discontinuous epitope across domains I and II. J Struct Biol X 12, 100135 (2025). 10.1016/j.yjsbx.2025.100135

12 Agar, C. et al. Beta2-glycoprotein I can exist in 2 conformations: implications for our understanding of the antiphospholipid syndrome. Blood 116, 1336–1343 (2010). 10.1182/blood-2009-12-260976

13 Kumar, S., Wulf, J., 2nd, Basore, K. & Pozzi, N. Structural analyses of beta(2)-glycoprotein I: is there a circular conformation? J Thromb Haemost 21, 3511–3521 (2023). 10.1016/j.jtha.2023.07.016

14 Muller-Calleja, N. et al. Antiphospholipid antibodies in a large population-based cohort: genome-wide associations and effects on monocyte gene expression. Thromb Haemost 116, 115–123 (2016). 10.1160/TH15-12-0947

15 Ghouse, J. et al. Genome-wide meta-analysis identifies 93 risk loci and enables risk prediction equivalent to monogenic forms of venous thromboembolism. Nat Genet 55, 399–409 (2023). 10.1038/s41588-022-01286-7

16 Zuber, V. et al. Combining evidence from Mendelian randomization and colocalization: Review and comparison of approaches. Am J Hum Genet 109, 767–782 (2022). 10.1016/j.ajhg.2022.04.001

17 Ng, P. C. & Henikoff, S. SIFT: Predicting amino acid changes that affect protein function. Nucleic Acids Res 31, 3812–3814 (2003). 10.1093/nar/gkg509

18 Reva, B., Antipin, Y. & Sander, C. Predicting the functional impact of protein mutations: application to cancer genomics. Nucleic Acids Res 39, e118 (2011). 10.1093/nar/gkr407

19 Choi, Y., Sims, G. E., Murphy, S., Miller, J. R. & Chan, A. P. Predicting the functional effect of amino acid substitutions and indels. PLoS One 7, e46688 (2012). 10.1371/journal.pone.0046688

20 Kircher, M. et al. A general framework for estimating the relative pathogenicity of human genetic variants. Nat Genet 46, 310–315 (2014). 10.1038/ng.2892

21 Quang, D., Chen, Y. & Xie, X. DANN: a deep learning approach for annotating the pathogenicity of genetic variants. Bioinformatics 31, 761–763 (2015). 10.1093/bioinformatics/btu703

22 Malhis, N., Jacobson, M., Jones, S. J. M. & Gsponer, J. LIST-S2: taxonomy based sorting of deleterious missense mutations across species. Nucleic Acids Res 48, W154–W161 (2020). 10.1093/nar/gkaa288

23 Rives, A. et al. Biological structure and function emerge from scaling unsupervised learning to 250 million protein sequences. Proc Natl Acad Sci U S A 118 (2021). 10.1073/pnas.2016239118

24 Minton, K. Predicting variant pathogenicity with AlphaMissense. Nat Rev Genet 24, 804 (2023). 10.1038/s41576-023-00668-9

25 Gao, H. et al. The landscape of tolerated genetic variation in humans and primates. bioRxiv (2023). 10.1101/2023.05.01.538953

26 Sanghera, D. K., Wagenknecht, D. R., McIntyre, J. A. & Kamboh, M. I. Identification of structural mutations in the fifth domain of apolipoprotein H (beta 2-glycoprotein I) which affect phospholipid binding. Hum Mol Genet 6, 311–316 (1997). 10.1093/hmg/6.2.311

27 Hasdemir, H. S., Pozzi, N. & Tajkhorshid, E. Atomistic characterization of beta2-glycoprotein I domain V interaction with anionic membranes. J Thromb Haemost 22, 3277–3289 (2024). 10.1016/j.jtha.2024.07.010

28 Kolyada, A., Barrios, D. A. & Beglova, N. Dimerized Domain V of Beta2-Glycoprotein I Is Sufficient to Upregulate Procoagulant Activity in PMA-Treated U937 Monocytes and Require Intact Residues in Two Phospholipid-Binding Loops. Antibodies (Basel) 6 (2017). 10.3390/antib6020008

29 Davies, N. M., Holmes, M. V. & Davey Smith, G. Reading Mendelian randomisation studies: a guide, glossary, and checklist for clinicians. BMJ 362, k601 (2018). 10.1136/bmj.k601

30 Chen, D. P. et al. PRTN3 variant correlates with increased autoantigen levels and relapse risk in PR3-ANCA versus MPO-ANCA disease. JCI Insight 8 (2023). 10.1172/jci.insight.166107

31 Xie, J. et al. The genetic architecture of membranous nephropathy and its potential to improve non-invasive diagnosis. Nat Commun 11, 1600 (2020). 10.1038/s41467-020-15383-w

32 Gschwind, A. R. et al. An encyclopedia of enhancer-gene regulatory interactions in the human genome. bioRxiv (2023). 10.1101/2023.11.09.563812

33 Mehdi, H., Aston, C. E., Sanghera, D. K., Hamman, R. F. & Kamboh, M. I. Genetic variation in the apolipoprotein H (beta2-glycoprotein I) gene affects plasma apolipoprotein H concentrations. Hum Genet 105, 63–71 (1999). 10.1007/s004399900089

34 Ruiu, G., Gambino, R., Veglia, F., Pagano, G. & Cassader, M. Influence of APOH protein polymorphism on apoH levels in normal and diabetic subjects. Clin Genet 52, 167–172 (1997). 10.1111/j.1399-0004.1997.tb02538.x

35 Hoie, M. H. et al. DiscoTope-3.0: improved B-cell epitope prediction using inverse folding latent representations. Front Immunol 15, 1322712 (2024). 10.3389/fimmu.2024.1322712

36 Dimitriadis, E., Menkhorst, E., Saito, S., Kutteh, W. H. & Brosens, J. J. Recurrent pregnancy loss. Nat Rev Dis Primers 6, 98 (2020). 10.1038/s41572-020-00228-z

37 Daniel J Wallace, D. D. G. UpToDate: Systemic lupus erythematosus in adults: Clinical manifestations and diagnosis, <https://www.uptodate.com/contents/systemic-lupus-erythematosus-in-adults-clinical-manifestations-and-diagnosis> (2025).

38 Casares-Marfil, D. et al. A Genome-Wide Association Study Suggests New Susceptibility Loci for Primary Antiphospholipid Syndrome. Arthritis Rheumatol 76, 1623–1634 (2024). 10.1002/art.42947

39 Giambartolomei, C. et al. Bayesian test for colocalisation between pairs of genetic association studies using summary statistics. PLoS Genet 10, e1004383 (2014). 10.1371/journal.pgen.1004383

40 Bild, D. E. et al. Multi-Ethnic Study of Atherosclerosis: objectives and design. Am J Epidemiol 156, 871–881 (2002). 10.1093/aje/kwf113

41 Fanopoulos, D., Teodorescu, M. R., Varga, J. & Teodorescu, M. High frequency of abnormal levels of IgA anti-beta2-glycoprotein I antibodies in patients with systemic lupus erythematosus: relationship with antiphospholipid syndrome. J Rheumatol 25, 675–680 (1998).

42 Manichaikul, A. et al. Genome-wide study of percent emphysema on computed tomography in the general population. The Multi-Ethnic Study of Atherosclerosis Lung/SNP Health Association Resource Study. Am J Respir Crit Care Med 189, 408–418 (2014). 10.1164/rccm.201306-1061OC

43 Genomes Project, C., et al. A global reference for human genetic variation. Nature 526, 68–74 (2015). 10.1038/nature15393

44 Patterson, N., Price, A. L. & Reich, D. Population structure and eigenanalysis. PLoS Genet 2, e190 (2006). 10.1371/journal.pgen.0020190

45 Das, S. et al. Next-generation genotype imputation service and methods. Nat Genet 48, 1284–1287 (2016). 10.1038/ng.3656

46 Taliun, D. et al. Sequencing of 53,831 diverse genomes from the NHLBI TOPMed Program. Nature 590, 290–299 (2021). 10.1038/s41586-021-03205-y

47 Loh, P. R., Palamara, P. F. & Price, A. L. Fast and accurate long-range phasing in a UK Biobank cohort. Nat Genet 48, 811–816 (2016). 10.1038/ng.3571

48 Das, S., Abecasis, G. R. & Browning, B. L. Genotype Imputation from Large Reference Panels. Annu Rev Genomics Hum Genet 19, 73–96 (2018). 10.1146/annurev-genom-083117-021602

49 Mbatchou, J. et al. Computationally efficient whole-genome regression for quantitative and binary traits. Nat Genet 53, 1097–1103 (2021). 10.1038/s41588-021-00870-7

50 Willer, C. J., Li, Y. & Abecasis, G. R. METAL: fast and efficient meta-analysis of genomewide association scans. Bioinformatics 26, 2190–2191 (2010). 10.1093/bioinformatics/btq340

51 Wang, G., Sarkar, A., Carbonetto, P. & Stephens, M. A simple new approach to variable selection in regression, with application to genetic fine mapping. J R Stat Soc Series B Stat Methodol 82, 1273–1300 (2020). 10.1111/rssb.12388

52 Gao, B. & Zhou, X. MESuSiE enables scalable and powerful multi-ancestry fine-mapping of causal variants in genome-wide association studies. Nat Genet 56, 170–179 (2024). 10.1038/s41588-023-01604-7

53 Wallace, C. A more accurate method for colocalisation analysis allowing for multiple causal variants. PLoS Genet 17, e1009440 (2021). 10.1371/journal.pgen.1009440

54 Consortium, E. P. et al. Expanded encyclopaedias of DNA elements in the human and mouse genomes. Nature 583, 699–710 (2020). 10.1038/s41586-020-2493-4

55 Roadmap Epigenomics, C., et al. Integrative analysis of 111 reference human epigenomes. Nature 518, 317–330 (2015). 10.1038/nature14248

56 Consortium, G. T. The GTEx Consortium atlas of genetic regulatory effects across human tissues. Science 369, 1318–1330 (2020). 10.1126/science.aaz1776

57 Sun, B. B. et al. Plasma proteomic associations with genetics and health in the UK Biobank. Nature 622, 329–338 (2023). 10.1038/s41586-023-06592-6

58 Landrum, M. J. et al. ClinVar: public archive of relationships among sequence variation and human phenotype. Nucleic Acids Res 42, D980–985 (2014). 10.1093/nar/gkt1113

59 Stenson, P. D. et al. The Human Gene Mutation Database (HGMD((R))): optimizing its use in a clinical diagnostic or research setting. Hum Genet 139, 1197–1207 (2020). 10.1007/s00439-020-02199-3

60 Kondo, A. et al. Glycopeptide profiling of beta-2-glycoprotein I by mass spectrometry reveals attenuated sialylation in patients with antiphospholipid syndrome. J Proteomics 73, 123–133 (2009). 10.1016/j.jprot.2009.08.007

61 Jo, S., Kim, T., Iyer, V. G. & Im, W. CHARMM-GUI: a web-based graphical user interface for CHARMM. J Comput Chem 29, 1859–1865 (2008). 10.1002/jcc.20945

62 Baerenfaenger, M. & Meyer, B. Simultaneous characterization of SNPs and N-glycans from multiple glycosylation sites of intact beta-2-glycoprotein-1 (B2GP1) by ESI-qTOF-MS. Biochim Biophys Acta Proteins Proteom 1867, 556–564 (2019). 10.1016/j.bbapap.2019.03.007

63 Phillips, J. C. et al. Scalable molecular dynamics on CPU and GPU architectures with NAMD. J Chem Phys 153, 044130 (2020). 10.1063/5.0014475

64 MacKerell, A. D. et al. All-atom empirical potential for molecular modeling and dynamics studies of proteins. J Phys Chem B 102, 3586–3616 (1998). 10.1021/jp973084f

65 Huang, J. & MacKerell, A. D., Jr. CHARMM36 all-atom additive protein force field: validation based on comparison to NMR data. J Comput Chem 34, 2135–2145 (2013). 10.1002/jcc.23354

66 Mackerell, A. D., Jr., Feig, M. & Brooks, C. L., 3rd. Extending the treatment of backbone energetics in protein force fields: limitations of gas-phase quantum mechanics in reproducing protein conformational distributions in molecular dynamics simulations. J Comput Chem 25, 1400–1415 (2004). 10.1002/jcc.20065

67 Glenn J. Martyna, D. J. T., Michael L. Klein. Constant pressure molecular dynamics algorithms The Journal of Chemical Physics 101, 4177–4189 (1994).

68 Scott E. Feller, Y. Z., Richard W. Pastor, Bernard R. Brooks. Constant pressure molecular dynamics simulation: The Langevin piston method The Journal of Chemical Physics 103, 4613–4621 (1995).

69 Grant, B. J., Rodrigues, A. P., ElSawy, K. M., McCammon, J. A. & Caves, L. S. Bio3d: an R package for the comparative analysis of protein structures. Bioinformatics 22, 2695–2696 (2006). 10.1093/bioinformatics/btl461

70 Lalaurie, C. J. et al. Elucidation of critical pH-dependent structural changes in Botulinum Neurotoxin E. J Struct Biol 214, 107876 (2022). 10.1016/j.jsb.2022.107876

71 Lalaurie, C. J., Zhang, C., Liu, S. M., Bunting, K. A. & Dalby, P. A. An open source in silico workflow to assist in the design of fusion proteins. Comput Biol Chem 113, 108209 (2024). 10.1016/j.compbiolchem.2024.108209

72 Spiteri, V. A. et al. Solution structure of deglycosylated human IgG1 shows the role of C(H)2 glycans in its conformation. Biophys J 120, 1814–1834 (2021). 10.1016/j.bpj.2021.02.038

73 Spiteri, V. A. et al. Solution structures of human myeloma IgG3 antibody reveal extended Fab and Fc regions relative to the other IgG subclasses. J Biol Chem 297, 100995 (2021). 10.1016/j.jbc.2021.100995

74 Humphrey, W., Dalke, A. & Schulten, K. VMD: visual molecular dynamics. J Mol Graph 14, 33–38, 27–38 (1996). 10.1016/0263-7855(96)00018-5

75 Chen, X. & Deng, Y. Long-time molecular dynamics simulations of botulinum biotoxin type-A at different pH values and temperatures. J Mol Model 13, 559–572 (2007). 10.1007/s00894-007-0178-7

76 Palm, K. et al. Evaluation of dynamic polar molecular surface area as predictor of drug absorption: comparison with other computational and experimental predictors. J Med Chem 41, 5382–5392 (1998). 10.1021/jm980313t

