## Supplementary Material for "A Common Missense Variant, W335S, in β2-Glycoprotein I (APOH) is Associated with Increased Autoantibody Levels but Reduced Venous Thromboembolism Risk"

**Supplementary Note**

**Evaluation of non-coding variants from the credible sets**

To assess overlap of candidate variants with regulatory regions inferred from epigenomic datasets, we obtained data from the ENCODE^1^ and Roadmap Epigenomics Projects^2^. For ENCODE, liver-specific candidate cis-Regulatory Elements (cCREs) annotations were obtained from the Registry of cCREs (Version 4). The identification of cCREs in ENCODE is primarily based on representative DNase I hypersensitivity sites (rDHSs), but also includes non-rDHS transcription factor binding sites that are reproducible across cell types, contain high-quality binding motifs, and are evolutionarily conserved. For the Roadmap Epigenomics Project , we obtained liver-specific annotations for H3K4ma1 (NarrowPeak and GappedPeak), H3K27ac (NarrowPeak and GappedPeak), H3K4m23 (NarrowPeak), and H3k9ac (NarrowPeak) from the consolidated chromatin immunoprecipitation followed by sequencing (ChIP-seq)-seq datasets from the Human Epigenome Atlas (Release 9).

To evaluate whether candidate variants overlap with elements predicted to regulate β2GPI (APOH) expression through cis-regulatory mechanisms, we obtained liver-specific annotations from ENCODE-rE2G.^3^ ENCODE-rE2G builds prediction models of enhancer–promoter interactions based on chromatin state features and 3D chromatin contacts, trained using CRISPR perturbation data. These models were then applied across 352 samples with DNase-seq data generated by the ENCODE Project. We extracted ENCODE-rE2G predictions from liver-related samples (“liver,” “left_lobe_of_liver,” “right_lobe_of_liver,” “hepatocyte,” “HepG2,” and “HuH-7.5”) and assessed overlap between candidate variants and elements predicted to regulate β2GPI (APOH) expression.

To assess the effect of candidate variants on gene expression, we obtained cis-regulatory summary statistics for the β2GPI (APOH) liver expression quantitative trait locus (eQTL) from the Genotype-Tissue Expression (GTEx, version 8).^4^ GTEx is a large-scale resource that characterizes cis-regulation of mRNA expression across diverse human tissues; the liver dataset includes 208 donor samples. We examined the effects of SNPs in the 95% credible sets for anti-β2GPI antibody levels at the APOH locus on β2GPI (APOH) expression and then performed colocalization analysis between anti-β2GPI antibody levels and Β2GPI (APOH) expression. Genotype data from European individuals in the 1000 Genomes Project were used as the LD reference panel for analyses of the GTEx data. In parallel, we also performed colocalization analysis between VTE and liver β2GPI (APOH) expression.

To assess the relationship between genetic associations for anti-β2GPI antibody levels and β2GPI (APOH) antigen levels, we obtained summary statistics for the Β2GPI (APOH) protein quantitative trait locus (pQTL) from the UK Biobank Pharma Proteomics Project (UKB-PPP).^5^

The UKB-PPP is a precompetitive biopharmaceutical consortium that mapped pQTLs for 2,923 plasma proteins in 54,219 UK Biobank participants using the antibody-based Olink Explore 3072 platform. We examined the effects of SNPs in the 95% credible sets for anti-β2GPI antibody levels at the *APOH* locus on Β2GPI (APOH) protein levels, and then performed colocalization analysis between anti-β2GPI antibody levels and plasma Β2GPI (APOH) protein levels. In-sample genotypes from the respective datasets were used as the LD reference panels. In parallel, we also performed colocalization analysis between VTE and plasma β2GPI (APOH) protein levels.

**Evaluation of protein coding variants from the credible sets**

To evaluate the functional impact of protein-coding variants, we applied ten prediction tools: SIFT^6^, MutationAssessor^7^, PROVEAN^8^, CADD^9^, DANN^10^, LIST-S2^11^, ESM-1b^12^, AlphaMissense^13^, and PrimateAI-3D^14^. Detailed descriptions of the selected prediction tools are provided in the below table. β2GPI

| **Tool** | **Version** | **Features used for model building** | **Brief method description** | **Reference dataset** | **Publication year** |
| --- | --- | --- | --- | --- | --- |
| SIFT (Sorting Intolerant From Tolerant) | Ensembl 66 | Position-specific substitution probabilities | Alignment-based method that predicts substitution impact by assessing conservation at the variant position using normalized probabilities | SWISS-PROT/TrEMBL and NCBI non-redundant protein databases | 2003^6^ |
| MutationAssessor | Release 3 | Conservation and specificity scores | Entropy-based scoring combining global conservation and subfamily-specific patterns to assess mutation impact | UniProtKB | 2011^7^ |
| PROVEAN (Protein Variation Effect Analyzer) | 1.1 | Delta alignment scores based on sequence similarity changes | Alignment-based delta score calculation measuring variant-induced changes in similarity to clustered homologous sequences | NCBI non-redundant (NR) protein database | 2012^8^ |
| CADD (Combined Annotation Dependent Depletion) | v1.7 | Diverse annotations including sequence properties, gene models, protein effects | Support vector machine distinguishing observed human variants from simulated deleterious ones | 14.7M high-frequency human-derived alleles vs. 14.7M simulated variants | 2014^9^ |
| DANN (Deep Neural Network) | NA | same as CADD | Deep neural network distinguishing observed human variants from simulated deleterious ones | Same as CADD | 2015^10^ |
| LIST-S2 (Local Identity and Taxonomy-based Sorting, v2) | V1.10 | Conservation measures based on local sequence identity and shared taxonomy with homologous sequences, and an amino acid swap-ability matrix (AASM) | Taxonomy-aware method quantifying conservation using local identity and taxonomic distances for cross-species predictions | UniProt for homology search. ExAC/gnomAD for allele frequencies | 2020^11^ |
| ESM-1b (Evolutionary Scale Modeling, 1b) | NA | Amino acid sequences | Protein language model; zero-shot variant effect via log-likelihood ratio (LLR) wild type vs mutant | ~250M protein sequences from UniProt/UniRef50 | 2021^12^ |
| AlphaMissense | NA | AlphaFold-based protein structures, MSA,and protein sequence context | Adapted AlphaFold model with protein language modeling, followed by fine-tunning on population frequency data | Large, diverse protein sequence databases. Human/primate population allele frequency databases for fine-tunnning | 2023^13^ |
| PrimateAI-3D | NA | AlphaFold-based voxelized 3D protein structures, multiple sequence alignments (MSAs), and common primate missense variants | Semi-supervised 3D-convolutional neural network | ~4.5M common missense variants from 233 primate species; structures from AlphaFold DB; MSAs for sequence context. | 2023^14^ |

**Sensitivity analysis**

Because rs1801690 had a minor allele frequency (MAF) of 0.0099 in African (AFR) individuals—slightly below the MAF ≥ 0.01 threshold—we performed a sensitivity analysis at this locus by including ancestry-specific variants with MAF ≥ 0.009. The multi-ancestry GWAS meta-analysis again identified rs1801690 as the lead SNP (p = 1.42 × 10⁻¹⁰). Fine-mapping using SuSiE under this threshold yielded results consistent with the primary analysis. SuSiE identified a single 95% credible set comprising 19 variants. The GWAS lead SNP, rs1801690, had the highest posterior inclusion probability (PIP = 0.44), whereas all other variants in the credible set showed much lower PIPs (<0.08).

At the *APOH* locus, There was evidence of colocalization between positive anti-β2GPI IgG (90th percentile cutoff) and VTE (PP4 = 0.92). Colocalization was suggestive between anti-β2GPI IgM (manufacturer’s cutoff) and VTE (PP4 = 0.89), as well as between anti-β2GPI IgM (90th percentile cutoff) and VTE (PP4 = 0.52).

We also performed two-sample MR using positive anti-β2GPI IgG (99th percentile cutoff) from an independent GWAS of 4,163 German individuals as the exposure, the reported lead SNP in this dataset (rs8178848) as the IV, and VTE as the outcome. rs8178848 was a strong instrument for positive anti-β2GPI IgG (99th percentile cutoff) in this external dataset (F-statistic = 46). Genetically determined positive anti-β2GPI IgG at the *APOH* locus was again negatively associated with VTE (β = –0.02, p = 5.3 x 10^-4^).

**Supplementary Table S1.** Ancestry-specific lead SNPs and corresponding rs1801690-G association statistics at the *APOH* locus for total anti-β2GPI antibody levels.

| Ancestral group | Lead SNP | MAF | Beta | P-value | Meta-analysis lead SNP | MAF | Beta | P-value |
| --- | --- | --- | --- | --- | --- | --- | --- | --- |
| EUR | rs9906486-T | 0.0527 | 0.23 | 1.59E-07 | rs1801690-G | 0.0529 | 0.22 | 2.90E-07 |
| AFR | rs74990880-T | 0.0265 | 0.22 | 5.00E-04 |  | 0.0099* | 0.09 | 3.94E-01 |
| AMR | rs2109910-G | 0.4208 | -0.07 | 4.90E-03 |  | 0.0193 | 0.24 | 5.98E-03 |
| EAS | rs148235559-T | 0.0262 | -0.27 | 1.14E-03 |  | 0.0614 | 0.17 | 3.50E-03 |

*Not included in the primary analysis (MAF cutoff 0.01); included in sensitivity analyses using MAF ≥ 0.009.

EUR: European; AFR: African; AMR: Admixed American; EAS: East Asian; MAF: minor allele frequency.

**Supplementary Table S2.** Association results at the *APOH* locus for anti-β2GPI antibody isotypes across different positivity thresholds.

| Isotype | Positivity threshold | Case/control numbers | Isotype-specific lead SNP | MAF | Beta | P-value | Total anti-β2GPI lead SNP | MAF | Beta | P-value |
| --- | --- | --- | --- | --- | --- | --- | --- | --- | --- | --- |
| IgM | **90th percentile** | **598/5128** | **rs7211380-G** | **0.06** | **0.72** | **8.44E-10** | rs1801690-G | **0.05** | **0.70** | **1.41E-06** |
|  | **95th percentile** | **286/5440** | **rs7211380-G** | **0.06** | **0.85** | **7.25E-09** |  | **0.05** | **0.89** | **9.08E-07** |
|  | 98th percentile | 117/5609 | rs114323586-T | 0.01 | 2.15 | 1.21E-04 |  | 0.05 | 0.22 | 5.75E-01 |
|  | 99th percentile | 60/5666 | rs8178836-T | 0.02 | 1.67 | 1.89E-04 |  | 0.05 | -0.03 | 9.63E-01 |
|  | **Manufacturer cutoff** | **498/5228** | **rs7211380-G** | **0.06** | **0.76** | **5.96E-10** |  | **0.05** | **0.76** | **5.23E-07** |
| IgG | **90th percentile** | **563/5163** | **rs11651658-C** | **0.06** | **0.69** | **3.34E-08** |  | **0.05** | **0.78** | **6.92E-07** |
|  | 95th percentile | 281/5445 | rs7211380-G | 0.06 | 0.70 | 1.07E-05 |  | 0.05 | 0.73 | 3.87E-04 |
|  | 98th percentile | 115/5611 | rs11651658-C | 0.07 | 0.95 | 7.50E-05 |  | 0.05 | 0.88 | 1.71E-03 |
|  | 99th percentile | 57/5669 | rs16959049-G | 0.03 | 1.71 | 6.78E-04 |  | 0.04 | 0.41 | 5.23E-01 |
|  | Manufacturer cutoff | 96/5630 | rs77048481-G | 0.03 | 1.58 | 3.41E-04 |  | 0.04 | 0.84 | 8.53E-02 |
| IgA | 90th percentile | 557/5169 | rs76375367-A | 0.06 | 0.55 | 1.48E-04 |  | 0.05 | 0.50 | 3.72E-03 |
|  | 95th percentile | 281/5445 | rs8077833-C | 0.04 | 1.07 | 9.06E-06 |  | 0.04 | 0.33 | 2.74E-01 |
|  | 98th percentile | 115/5611 | rs227907-G | 0.04 | 1.44 | 3.51E-03 |  | 0.04 | 0.41 | 3.86E-01 |
|  | 99th percentile | 57/5669 | rs182445372-C | 0.06 | 0.99 | 1.58E-02 |  | 0.02 | -0.30 | 7.62E-01 |
|  | Manufacturer cutoff | 565/5161 | rs76375367-A | 0.06 | 0.52 | 3.30E-04 |  | 0.05 | 0.47 | 6.10E-03 |

*Binary trait GWAS performed only in unrelated individuals, as the REGENIE LOCO scheme was unstable with small case numbers. Genome-wide significant loci (p < 5 × 10⁻⁸) are shown in bold. MAF: minor allele frequency.

**Supplementary Table S3.** SNPs included in the 95% credible set identified by SuSiE at the *APOH* locus associated with total anti-β2GPI antibody levels.

| SNP | CHR | BP | A1 | A2 | Het I^2^ | Het P | SuSiE PIP | Population | MAF | Beta | SE | P |
| --- | --- | --- | --- | --- | --- | --- | --- | --- | --- | --- | --- | --- |
| rs1801690 | 17 | 66212167 | G | C | 0 | 0.70 | 0.49 | Meta-Analysis | 0.05 | 0.21 | 0.03 | 1.08E-10 |
|  |  |  |  |  |  |  |  | EUR | 0.05 | 0.22 | 0.04 | 2.90E-07 |
|  |  |  |  |  |  |  |  | AFR | 0.01 | 0.09 | 0.10 | 3.94E-01 |
|  |  |  |  |  |  |  |  | AMR | 0.02 | 0.24 | 0.09 | 5.98E-03 |
|  |  |  |  |  |  |  |  | EAS | 0.06 | 0.17 | 0.06 | 3.50E-03 |
| rs9902706 | 17 | 66190870 | T | C | 47 | 0.13 | 0.06 | Meta-Analysis | 0.06 | 0.15 | 0.02 | 1.01E-09 |
|  |  |  |  |  |  |  |  | EUR | 0.05 | 0.22 | 0.04 | 2.54E-07 |
|  |  |  |  |  |  |  |  | AFR | 0.07 | 0.08 | 0.04 | 3.75E-02 |
|  |  |  |  |  |  |  |  | AMR | 0.03 | 0.19 | 0.08 | 1.28E-02 |
|  |  |  |  |  |  |  |  | EAS | 0.06 | 0.14 | 0.06 | 1.56E-02 |
| rs9906486 | 17 | 66194802 | T | G | 55 | 0.08 | 0.05 | Meta-Analysis | 0.06 | 0.15 | 0.02 | 1.31E-09 |
|  |  |  |  |  |  |  |  | EUR | 0.05 | 0.23 | 0.04 | 1.59E-07 |
|  |  |  |  |  |  |  |  | AFR | 0.07 | 0.08 | 0.04 | 6.27E-02 |
|  |  |  |  |  |  |  |  | AMR | 0.03 | 0.18 | 0.07 | 1.32E-02 |
|  |  |  |  |  |  |  |  | EAS | 0.06 | 0.14 | 0.06 | 1.14E-02 |
| rs9905408 | 17 | 66194566 | C | A | 56 | 0.08 | 0.04 | Meta-Analysis | 0.06 | 0.15 | 0.02 | 1.52E-09 |
|  |  |  |  |  |  |  |  | EUR | 0.05 | 0.23 | 0.04 | 1.59E-07 |
|  |  |  |  |  |  |  |  | AFR | 0.07 | 0.07 | 0.04 | 7.18E-02 |
|  |  |  |  |  |  |  |  | AMR | 0.03 | 0.18 | 0.07 | 1.32E-02 |
|  |  |  |  |  |  |  |  | EAS | 0.06 | 0.14 | 0.06 | 1.14E-02 |
| rs7211380 | 17 | 66210650 | G | A | 0 | 0.46 | 0.04 | Meta-Analysis | 0.06 | 0.15 | 0.02 | 1.64E-09 |
|  |  |  |  |  |  |  |  | EUR | 0.07 | 0.18 | 0.04 | 2.13E-06 |
|  |  |  |  |  |  |  |  | AFR | 0.05 | 0.09 | 0.05 | 6.08E-02 |
|  |  |  |  |  |  |  |  | AMR | 0.03 | 0.15 | 0.07 | 2.52E-02 |
|  |  |  |  |  |  |  |  | EAS | 0.06 | 0.16 | 0.06 | 4.88E-03 |
| rs74934196 | 17 | 66154754 | A | AAC | 0 | 0.65 | 0.03 | Meta-Analysis | 0.05 | 0.18 | 0.03 | 1.98E-09 |
|  |  |  |  |  |  |  |  | EUR | 0.05 | 0.21 | 0.04 | 8.84E-07 |
|  |  |  |  |  |  |  |  | AFR | 0.01 | 0.11 | 0.10 | 2.83E-01 |
|  |  |  |  |  |  |  |  | AMR | 0.02 | 0.21 | 0.09 | 1.60E-02 |
|  |  |  |  |  |  |  |  | EAS | 0.06 | 0.15 | 0.06 | 1.07E-02 |
| rs74531840 | 17 | 66179997 | A | T | 0 | 0.51 | 0.03 | Meta-Analysis | 0.05 | 0.18 | 0.03 | 2.43E-09 |
|  |  |  |  |  |  |  |  | EUR | 0.05 | 0.22 | 0.04 | 4.81E-07 |
|  |  |  |  |  |  |  |  | AFR | 0.01 | 0.08 | 0.09 | 3.53E-01 |
|  |  |  |  |  |  |  |  | AMR | 0.02 | 0.19 | 0.08 | 2.17E-02 |
|  |  |  |  |  |  |  |  | EAS | 0.06 | 0.14 | 0.06 | 1.13E-02 |
| rs76375367 | 17 | 66142666 | A | G | 42 | 0.16 | 0.02 | Meta-Analysis | 0.06 | 0.15 | 0.02 | 2.94E-09 |
|  |  |  |  |  |  |  |  | EUR | 0.05 | 0.22 | 0.04 | 5.21E-07 |
|  |  |  |  |  |  |  |  | AFR | 0.07 | 0.08 | 0.04 | 4.31E-02 |
|  |  |  |  |  |  |  |  | AMR | 0.03 | 0.15 | 0.07 | 4.52E-02 |
|  |  |  |  |  |  |  |  | EAS | 0.07 | 0.15 | 0.06 | 7.83E-03 |
| rs55657678 | 17 | 66144550 | A | G | 42 | 0.16 | 0.02 | Meta-Analysis | 0.06 | 0.15 | 0.02 | 2.73E-09 |
|  |  |  |  |  |  |  |  | EUR | 0.05 | 0.22 | 0.04 | 5.21E-07 |
|  |  |  |  |  |  |  |  | AFR | 0.07 | 0.08 | 0.04 | 4.31E-02 |
|  |  |  |  |  |  |  |  | AMR | 0.03 | 0.15 | 0.07 | 4.84E-02 |
|  |  |  |  |  |  |  |  | EAS | 0.06 | 0.16 | 0.06 | 6.71E-03 |
| rs11651658 | 17 | 66202522 | C | T | 0 | 0.57 | 0.02 | Meta-Analysis | 0.06 | 0.14 | 0.02 | 3.32E-09 |
|  |  |  |  |  |  |  |  | EUR | 0.07 | 0.17 | 0.04 | 4.45E-06 |
|  |  |  |  |  |  |  |  | AFR | 0.05 | 0.09 | 0.05 | 5.72E-02 |
|  |  |  |  |  |  |  |  | AMR | 0.03 | 0.16 | 0.07 | 1.92E-02 |
|  |  |  |  |  |  |  |  | EAS | 0.07 | 0.14 | 0.06 | 8.83E-03 |
| rs73992250 | 17 | 66136049 | G | A | 42 | 0.16 | 0.02 | Meta-Analysis | 0.06 | 0.15 | 0.02 | 3.77E-09 |
|  |  |  |  |  |  |  |  | EUR | 0.05 | 0.22 | 0.04 | 4.75E-07 |
|  |  |  |  |  |  |  |  | AFR | 0.07 | 0.08 | 0.04 | 4.31E-02 |
|  |  |  |  |  |  |  |  | AMR | 0.03 | 0.15 | 0.07 | 4.52E-02 |
|  |  |  |  |  |  |  |  | EAS | 0.06 | 0.15 | 0.06 | 1.09E-02 |
| rs7216660 | 17 | 66151258 | T | C | 42 | 0.16 | 0.02 | Meta-Analysis | 0.06 | 0.15 | 0.02 | 3.55E-09 |
|  |  |  |  |  |  |  |  | EUR | 0.05 | 0.22 | 0.04 | 4.73E-07 |
|  |  |  |  |  |  |  |  | AFR | 0.07 | 0.08 | 0.04 | 4.14E-02 |
|  |  |  |  |  |  |  |  | AMR | 0.03 | 0.15 | 0.07 | 4.64E-02 |
|  |  |  |  |  |  |  |  | EAS | 0.06 | 0.15 | 0.06 | 1.07E-02 |
| rs9891968 | 17 | 66136728 | A | G | 42 | 0.16 | 0.02 | Meta-Analysis | 0.06 | 0.15 | 0.02 | 3.97E-09 |
|  |  |  |  |  |  |  |  | EUR | 0.05 | 0.22 | 0.04 | 5.21E-07 |
|  |  |  |  |  |  |  |  | AFR | 0.07 | 0.08 | 0.04 | 4.31E-02 |
|  |  |  |  |  |  |  |  | AMR | 0.03 | 0.15 | 0.07 | 4.52E-02 |
|  |  |  |  |  |  |  |  | EAS | 0.06 | 0.15 | 0.06 | 1.09E-02 |
| rs9908597 | 17 | 66138565 | G | T | 42 | 0.16 | 0.02 | Meta-Analysis | 0.06 | 0.15 | 0.02 | 3.97E-09 |
|  |  |  |  |  |  |  |  | EUR | 0.05 | 0.22 | 0.04 | 5.21E-07 |
|  |  |  |  |  |  |  |  | AFR | 0.07 | 0.08 | 0.04 | 4.31E-02 |
|  |  |  |  |  |  |  |  | AMR | 0.03 | 0.15 | 0.07 | 4.52E-02 |
|  |  |  |  |  |  |  |  | EAS | 0.06 | 0.15 | 0.06 | 1.09E-02 |
| rs9895407 | 17 | 66161737 | C | T | 41 | 0.16 | 0.02 | Meta-Analysis | 0.06 | 0.15 | 0.02 | 4.33E-09 |
|  |  |  |  |  |  |  |  | EUR | 0.05 | 0.22 | 0.04 | 4.73E-07 |
|  |  |  |  |  |  |  |  | AFR | 0.07 | 0.08 | 0.04 | 3.93E-02 |
|  |  |  |  |  |  |  |  | AMR | 0.03 | 0.15 | 0.07 | 4.61E-02 |
|  |  |  |  |  |  |  |  | EAS | 0.06 | 0.14 | 0.06 | 1.44E-02 |
| rs77620153 | 17 | 66134676 | A | G | 43 | 0.16 | 0.02 | Meta-Analysis | 0.06 | 0.15 | 0.02 | 4.62E-09 |
|  |  |  |  |  |  |  |  | EUR | 0.05 | 0.22 | 0.04 | 5.21E-07 |
|  |  |  |  |  |  |  |  | AFR | 0.07 | 0.08 | 0.04 | 4.82E-02 |
|  |  |  |  |  |  |  |  | AMR | 0.03 | 0.15 | 0.07 | 4.52E-02 |
|  |  |  |  |  |  |  |  | EAS | 0.06 | 0.15 | 0.06 | 1.08E-02 |
| rs73992258 | 17 | 66168965 | C | A | 41 | 0.16 | 0.02 | Meta-Analysis | 0.06 | 0.15 | 0.02 | 4.47E-09 |
|  |  |  |  |  |  |  |  | EUR | 0.05 | 0.22 | 0.04 | 4.80E-07 |
|  |  |  |  |  |  |  |  | AFR | 0.07 | 0.08 | 0.04 | 3.93E-02 |
|  |  |  |  |  |  |  |  | AMR | 0.03 | 0.15 | 0.07 | 4.64E-02 |
|  |  |  |  |  |  |  |  | EAS | 0.06 | 0.14 | 0.06 | 1.47E-02 |
| rs9911603 | 17 | 66135784 | T | C | 40 | 0.17 | 0.02 | Meta-Analysis | 0.06 | 0.15 | 0.02 | 4.60E-09 |
|  |  |  |  |  |  |  |  | EUR | 0.05 | 0.22 | 0.04 | 6.40E-07 |
|  |  |  |  |  |  |  |  | AFR | 0.07 | 0.08 | 0.04 | 4.31E-02 |
|  |  |  |  |  |  |  |  | AMR | 0.03 | 0.15 | 0.07 | 4.52E-02 |
|  |  |  |  |  |  |  |  | EAS | 0.06 | 0.15 | 0.06 | 1.09E-02 |

CHR: chromosome; BP: base pair position; A1: effect allele; A2: non-effect allele; Het: heterogeneity; PIP: posterior inclusion probability; MAF: minor allele frequency; SE: standard error; EUR: European; AFR: African; AMR: Admixed American; EAS: East Asian.

**Supplementary Table S4.** Posterior inclusion probabilities (PIP) of SNPs in the 95% credible set prioritized by MESuSiE. The table reports the total PIP as well as ancestry-specific PIP values for each configuration. For comparison, PIP values from SuSiE applied to the multi-ancestry meta-analysis summary statistics are also provided. (This table is supplied as a separate Excel file.)

**Supplementary Table S5.** Colocalization analysis of anti-β2GPI antibody levels, venous thromboembolism (VTE), liver β2GPI (APOH) mRNA expression, and plasma β2GPI (APOH) protein levels at the ***APOH*** locus. No evidence of colocalization was observed between anti-β2GPI antibody levels and liver β2GPI (APOH) mRNA expression, between anti-β2GPI antibody levels and plasma β2GPI (APOH) protein levels, between VTE and liver β2GPI (APOH) mRNA expression, or between VTE and plasma β2GPI (APOH) protein levels.

| **Anti-β2GPI antibody levels and liver β2GPI (APOH) mRNA expression** | | | |
| --- | --- | --- | --- |
| Lead SNP in anti-β2GPI | Lead SNP in β2GPI (APOH) | PP3 | PP4 |
| rs1801690 | rs112389044 | 98.3365% | 0.0813% |
| **Anti-β2GPI antibody levels and plasma β2GPI (APOH) protein levels** | | | |
| Lead SNP in anti-β2GPI | Lead SNP in Β2GPI (APOH) | PP3 | PP4 |
| rs1801690 | rs144579769 | 90.2086% | 9.7861% |
|  | rs117299270 | 99.9939% | 0.0001% |
|  | rs58835646 | 99.9938% | 0.0003% |
|  | rs60530184 | 99.9938% | 0.0003% |
|  | rs116879328 | 99.9939% | 0.0001% |
|  | rs73992176 | 99.9938% | 0.0003% |
|  | rs145706684 | 99.9938% | 0.0003% |
|  | rs9906319 | 89.4147% | 10.5800% |
|  | rs1801689 | 99.8835% | 0.1106% |
|  | rs117423095 | 99.9936% | 0.0004% |
| **VTE and liver β2GPI (APOH) mRNA expression** | | | |
| Lead SNP in VTE | Lead SNP in β2GPI (APOH) | PP3 | PP4 |
| rs73992267 | rs112389044 | 97.9690% | 0.1689% |
| **VTE and plasma β2GPI (APOH) protein levels** | | | |
| Lead SNP in VTE | Lead SNP in β2GPI (APOH) | PP3 | PP4 |
| rs73992267 | rs144579769 | 94.0509% | 5.6710% |
|  | rs117299270 | 99.6945% | 0.0109% |
|  | rs58835646 | 99.6952% | 0.0101% |
|  | rs60530184 | 99.6975% | 0.0079% |
|  | rs116879328 | 99.6942% | 0.0111% |
|  | rs73992176 | 99.6954% | 0.0099% |
|  | rs145706684 | 99.6961% | 0.0092% |
|  | rs9906319 | 92.8446% | 6.8809% |
|  | rs1801689 | 99.6850% | 0.0204% |
|  | rs117423095 | 99.3568% | 0.3495% |

* Lead SNP is the lead SNP (highest posterior inclusion probability) from the SuSiE credible set; PP3: posterior probability for hypothesis 3 (two distinct causal variants); PP4: posterior probability for hypothesis 4 (a shared causal variant); VTE: thromboembolism.

**Supplementary Table S6.** Variant effect predictions for rs1801690 (W335S) using nine bioinformatic tools. These tools were selected because they have well-established cutoffs and were not trained on datasets containing Mendelian disease labels, such as ClinVar or Human Gene Mutation Database (HGMD).

| **Tools** | **Score** | **Interpretation** |
| --- | --- | --- |
| SIFT | 0.022 | Deleterious |
| MutationAssessor | 2.52 | Medium deleterious probability |
| PROVEAN | -3.7 | Deleterious |
| CADD | 28.6 (phred-scaled) | Deleterious |
| DANN | 0.99 | Deleterious |
| LIST-S2 | 0.73 | Tolerated |
| ESM-1b | -8 | Deleterious |
| AlphaMissense | 0.86 | Pathogenic |
| PrimateAI-3D | 0.52 | Benign |

SIFT: Sorting Intolerant From Tolerant; PROVEAN: Protein Variation Effect Analyzer; CADD: Combined Annotation Dependent Depletion; DANN (Deep Neural Network); LIST-S2 (Local Identity and Taxonomy-based Sorting, v2); ESM-1b: Evolutionary Scale Modeling 1b.

**Figure legend**

**Supplementary Figure S1.** Principal component analysis (PCA) of the 5,969 participants included in the primary GWAS analysis. PCs from MESA participants were projected into the PCA space defined by individuals from the 1000 Genomes Project. Genetically inferred ancestries included 2,354 European (EUR), 1,572 African (AFR), 1,294 Admixed American (AMR), and 749 East Asian (EAS) individuals. Participants who were outliers in ancestry-specific PCA (using SmartPCA) were excluded.

**Supplementary Figure S2.** Manhattan and QQ plots from GWAS of anti-β2GPI IgM and IgG at the 90th percentile cutoff.

**Supplementary Figure S3.** Regional plots of the *APOH* region for each ancestry group with corresponding MESuSiE posterior inclusion probabilities (PIP). MESuSiE identified rs1801690 as the variant with the highest PIP (0.23). The signal was predicted to be shared across EUR, AMR, and EAS, but not AFR.

**Supplementary Figure S4.** Distribution of expected causal signals across ancestry configurations from MESuSiE. Bars show the proportion of the expected number of signals (ENS) assigned to each of 15 configurations. The total ENS was approximately 2.0, with most support for a shared AMR–EUR–EAS configuration and additional support for EUR-centric configurations.

**Supplementary Figure S5.** LD heatmap for the 18 SNPs in the 95% credible set identified by SuSiE at the APOH locus. The SNP with the highest posterior inclusion probability (PIP), rs1801690, is also the GWAS lead SNP. rs1801690 showed moderate-to-strong LD with the other SNPs in EUR, AMR, and EAS, but only weak LD in AFR.

**Supplementary Figure S6.** Regional plots of the APOH region for total anti-β2GPI antibody levels, liver β2GPI (APOH) mRNA expression and plasma β2GPI (APOH) protein levels. There was no evidence of colocalization between anti-β2GPI antibody and liver β2GPI (APOH) mRNA expression (PP4 < 0.01), or between anti-β2GPI antibody and plasma β2GPI (APOH) protein levels (maximum PP4 = 0.11). In the fine-mapping plots, SNPs within the 95% credible sets are colored blue, and those with PP4 > 0.4 are labeled. Note there was a substantial difference in sample size between the liver eQTL dataset (n = 208) and the plasma pQTL dataset (n = 51,031).

1	Consortium, E. P. *et al.* Expanded encyclopaedias of DNA elements in the human and mouse genomes. *Nature* **583**, 699–710 (2020). <https://doi.org/10.1038/s41586-020-2493-4>

2	Roadmap Epigenomics, C. *et al.* Integrative analysis of 111 reference human epigenomes. *Nature* **518**, 317–330 (2015). <https://doi.org/10.1038/nature14248>

3	Gschwind, A. R. *et al.* An encyclopedia of enhancer-gene regulatory interactions in the human genome. *bioRxiv* (2023). <https://doi.org/10.1101/2023.11.09.563812>

4	Consortium, G. T. The GTEx Consortium atlas of genetic regulatory effects across human tissues. *Science* **369**, 1318–1330 (2020). <https://doi.org/10.1126/science.aaz1776>

5	Sun, B. B. *et al.* Plasma proteomic associations with genetics and health in the UK Biobank. *Nature* **622**, 329–338 (2023). <https://doi.org/10.1038/s41586-023-06592-6>

6	Ng, P. C. & Henikoff, S. SIFT: Predicting amino acid changes that affect protein function. *Nucleic Acids Res* **31**, 3812–3814 (2003). <https://doi.org/10.1093/nar/gkg509>

7	Reva, B., Antipin, Y. & Sander, C. Predicting the functional impact of protein mutations: application to cancer genomics. *Nucleic Acids Res* **39**, e118 (2011). <https://doi.org/10.1093/nar/gkr407>

8	Choi, Y., Sims, G. E., Murphy, S., Miller, J. R. & Chan, A. P. Predicting the functional effect of amino acid substitutions and indels. *PLoS One* **7**, e46688 (2012). <https://doi.org/10.1371/journal.pone.0046688>

9	Kircher, M. *et al.* A general framework for estimating the relative pathogenicity of human genetic variants. *Nat Genet* **46**, 310–315 (2014). <https://doi.org/10.1038/ng.2892>

10	Quang, D., Chen, Y. & Xie, X. DANN: a deep learning approach for annotating the pathogenicity of genetic variants. *Bioinformatics* **31**, 761–763 (2015). <https://doi.org/10.1093/bioinformatics/btu703>

11	Malhis, N., Jacobson, M., Jones, S. J. M. & Gsponer, J. LIST-S2: taxonomy based sorting of deleterious missense mutations across species. *Nucleic Acids Res* **48**, W154–W161 (2020). <https://doi.org/10.1093/nar/gkaa288>

12	Rives, A. *et al.* Biological structure and function emerge from scaling unsupervised learning to 250 million protein sequences. *Proc Natl Acad Sci U S A* **118** (2021). <https://doi.org/10.1073/pnas.2016239118>

13	Minton, K. Predicting variant pathogenicity with AlphaMissense. *Nat Rev Genet* **24**, 804 (2023). <https://doi.org/10.1038/s41576-023-00668-9>

14	Gao, H. *et al.* The landscape of tolerated genetic variation in humans and primates. *bioRxiv* (2023). <https://doi.org/10.1101/2023.05.01.538953>
