## Supplementary figures and images for "A Common Missense Variant, W335S, in β2-Glycoprotein I (APOH) is Associated with Increased Autoantibody Levels but Reduced Venous Thromboembolism Risk"

### Supplementary Figure S1

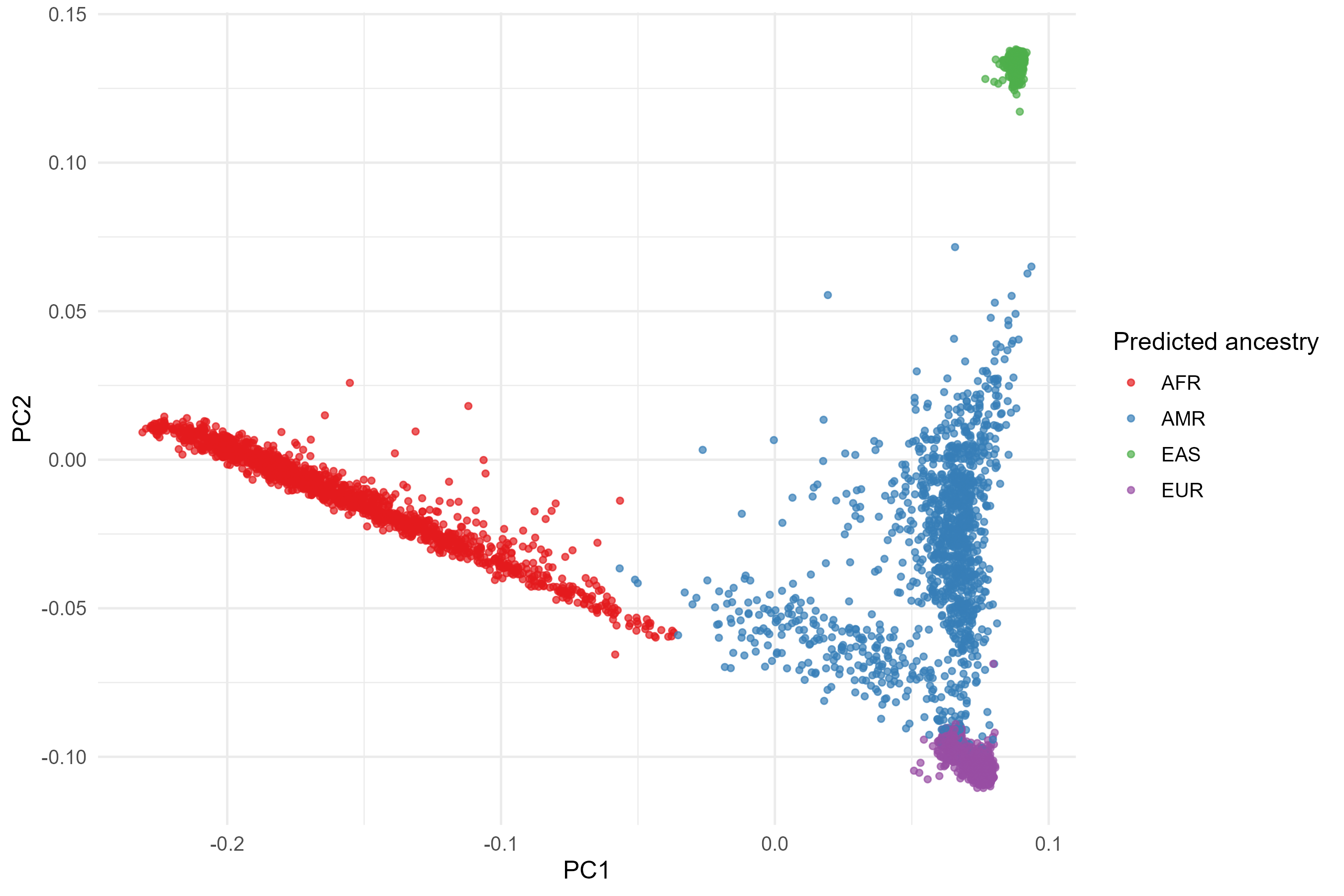

### Supplementary Figure S2

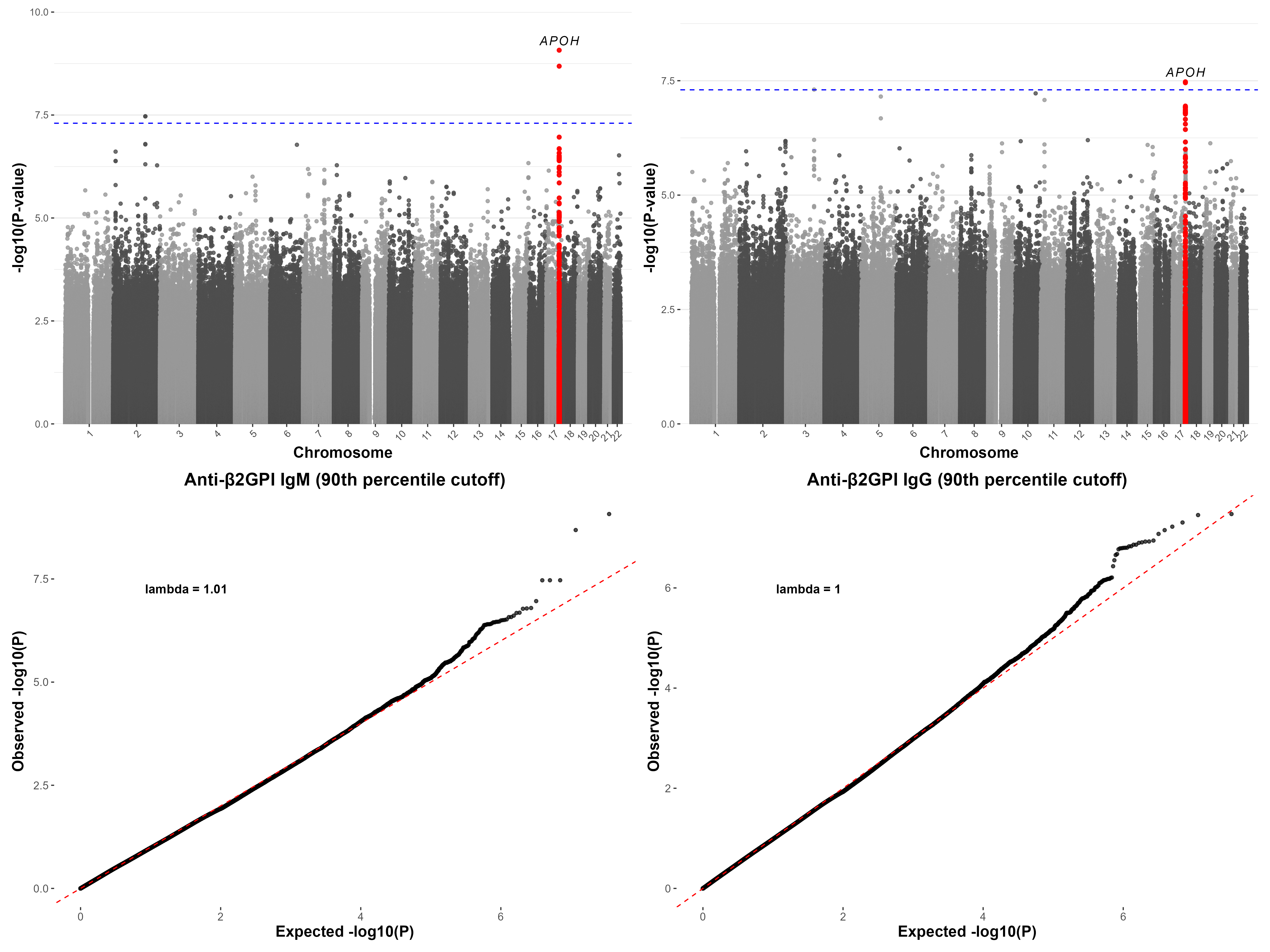

### Supplementary Figure S3

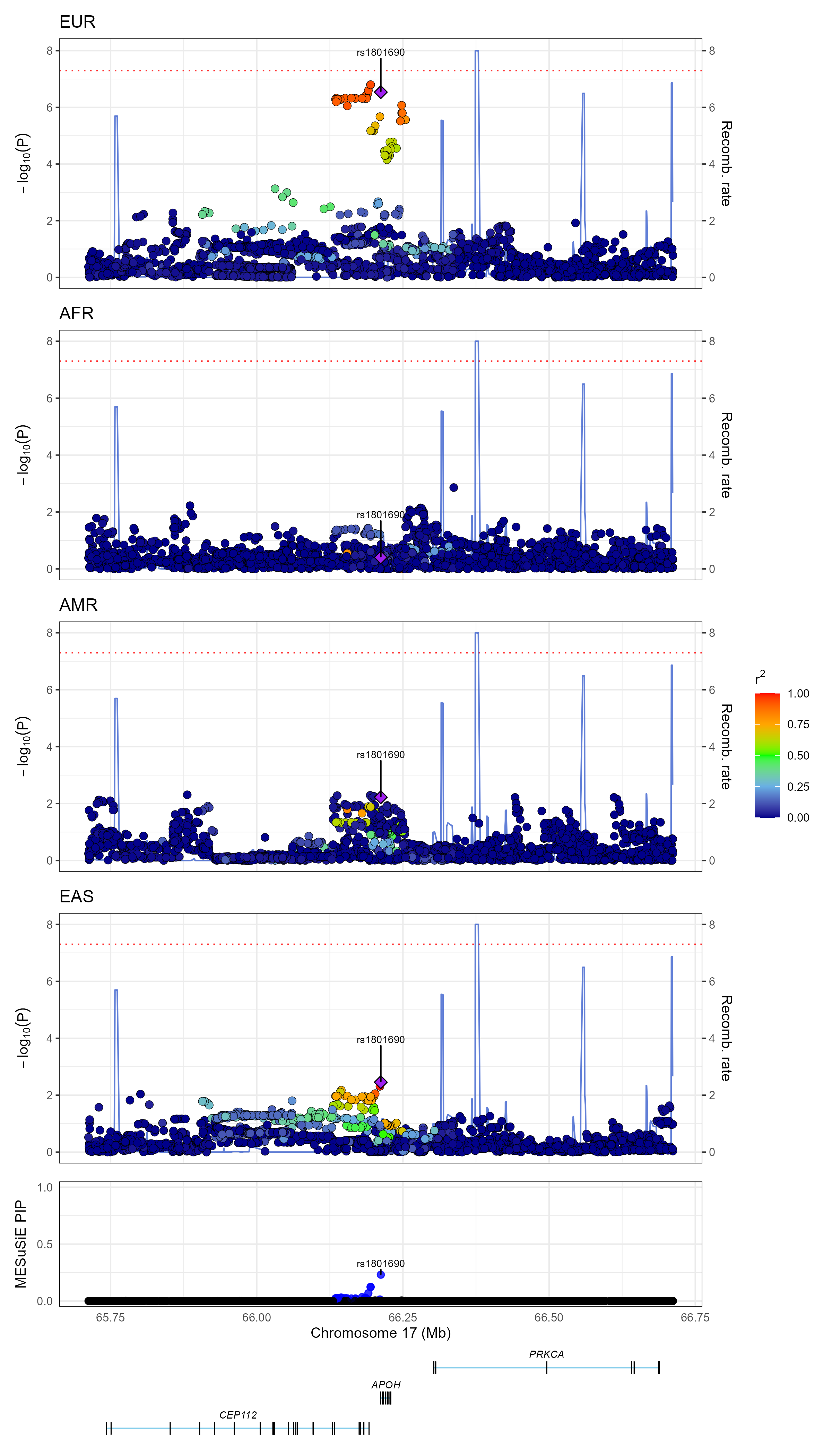

### Supplementary Figure S4

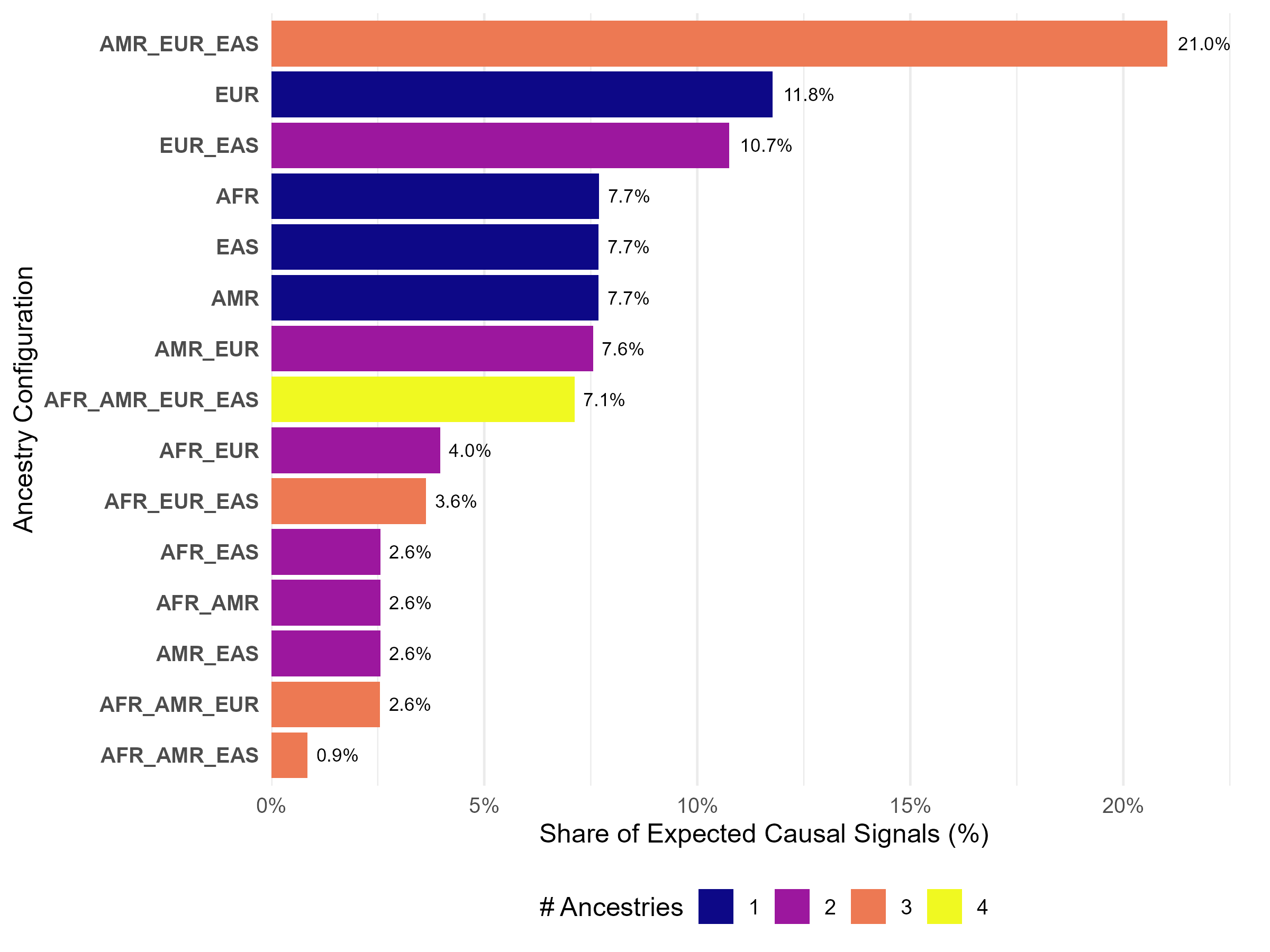

### Supplementary Figure S5

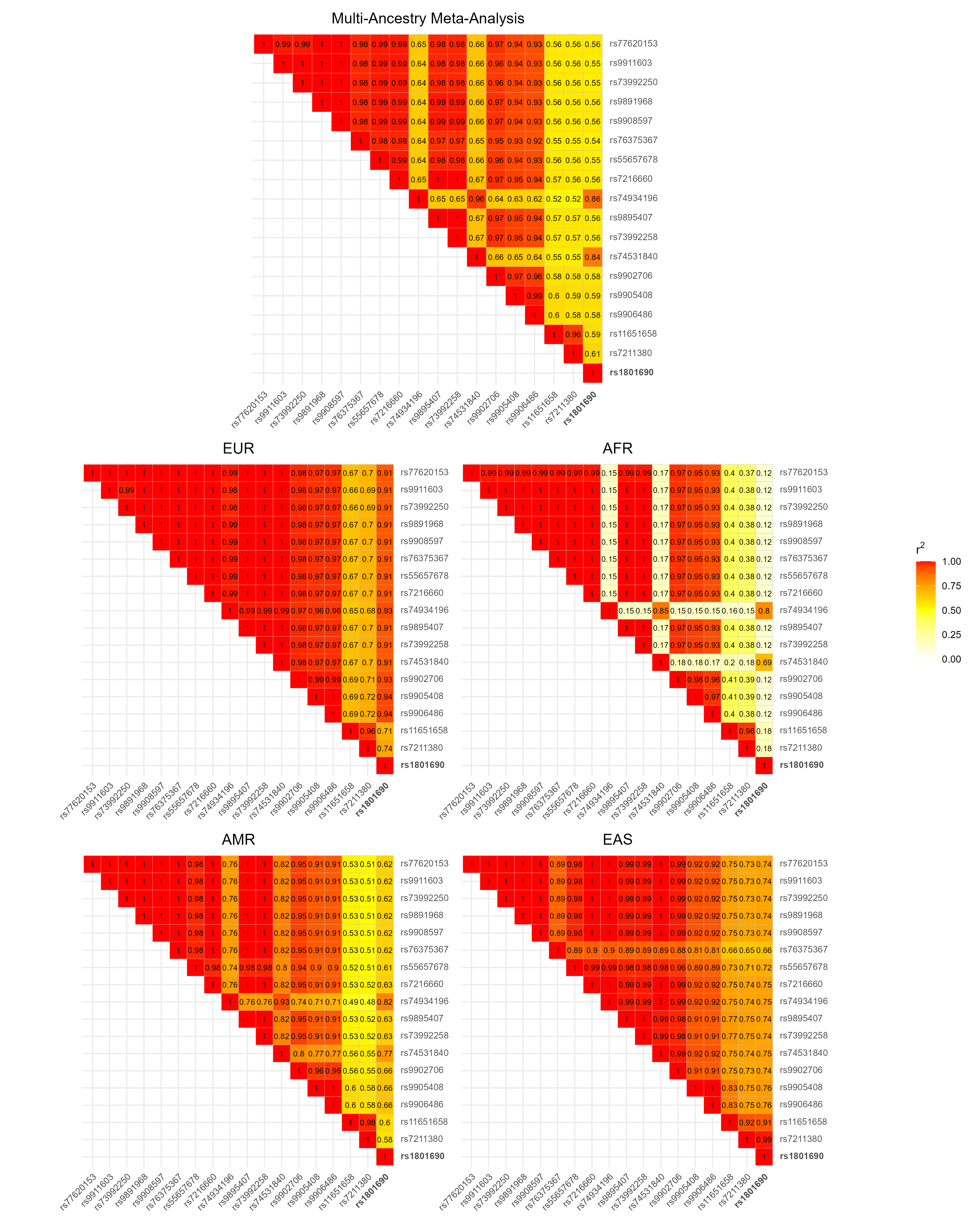

### Supplementary Figure S6

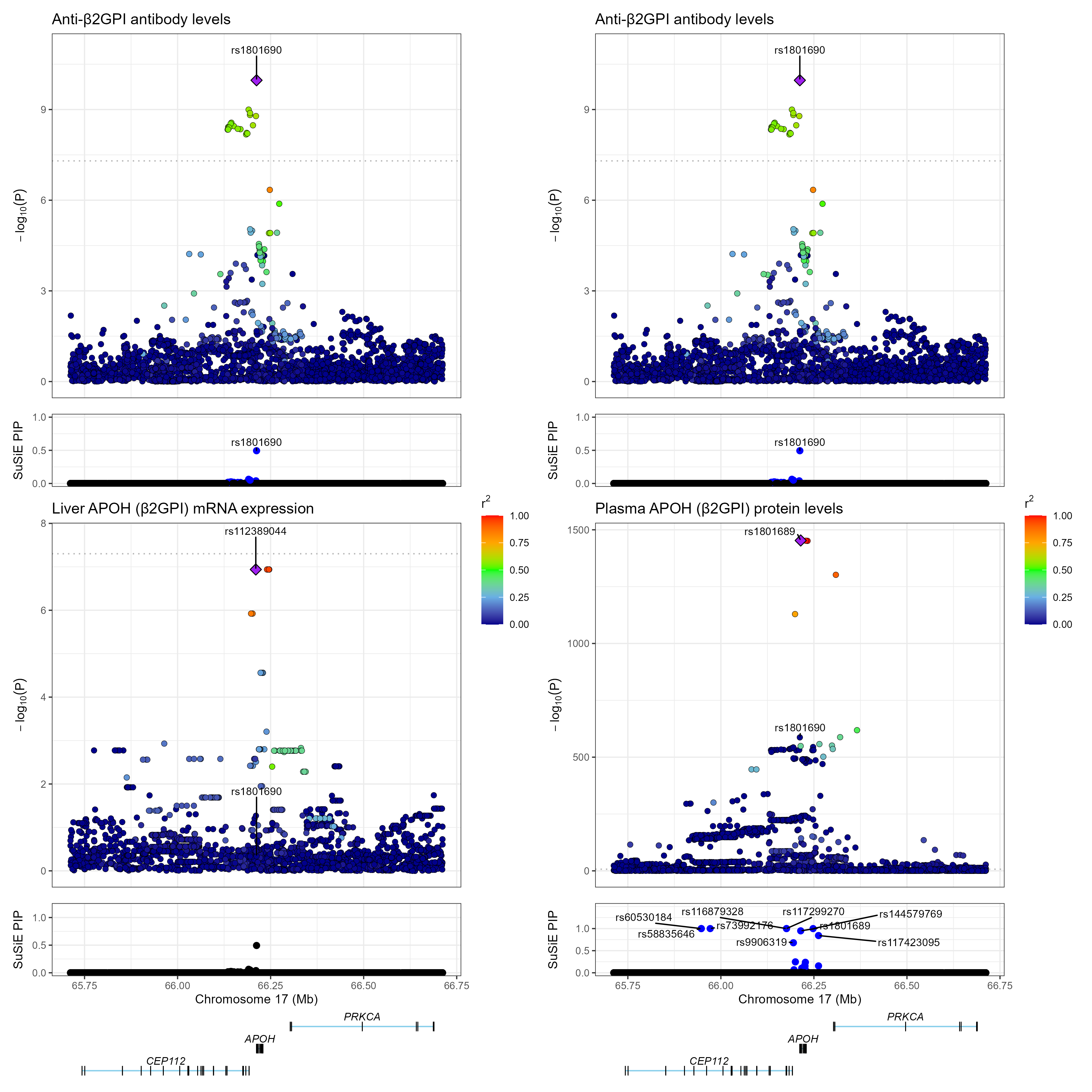

### Supplementary Figure S7

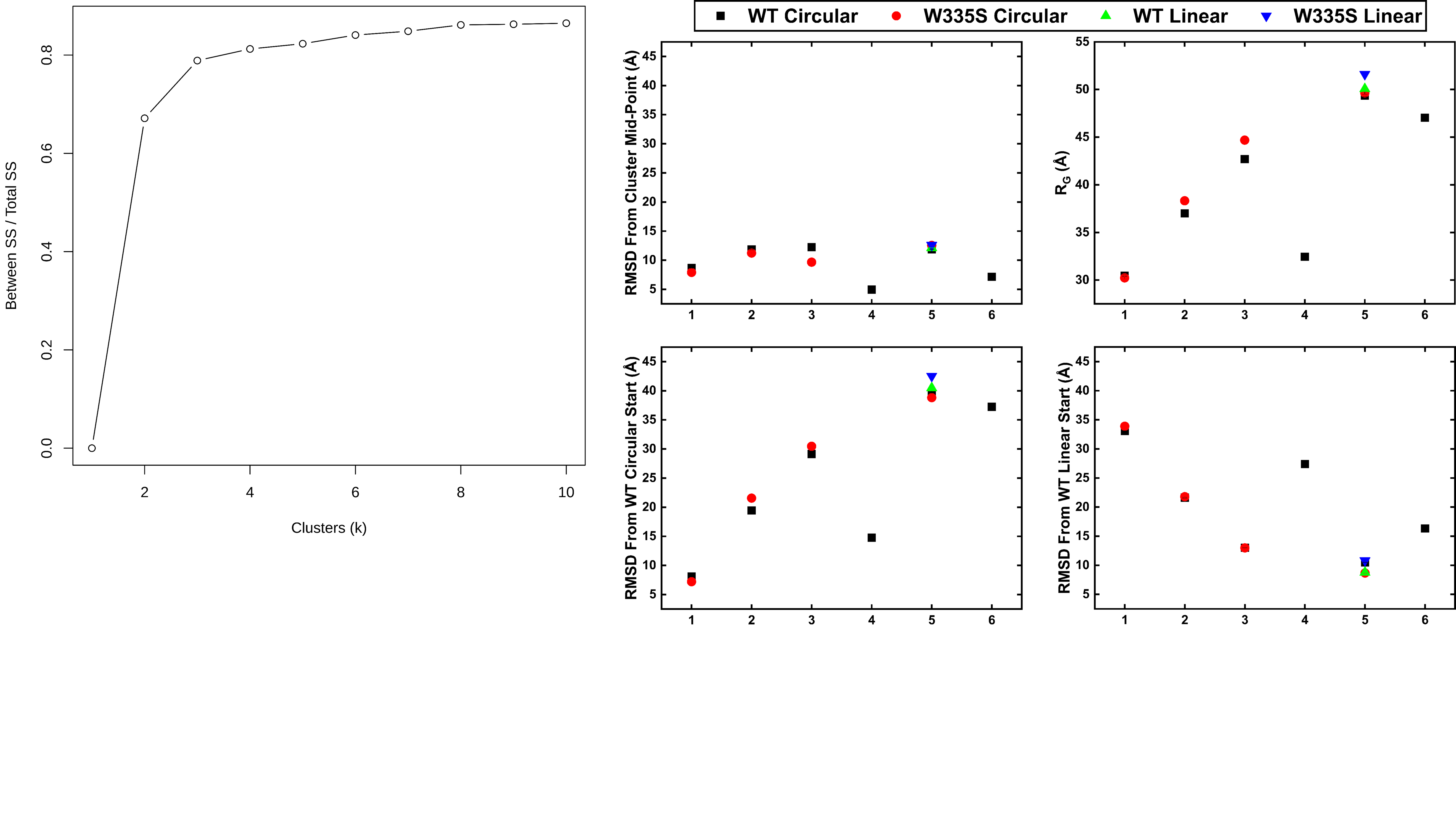

### Supplementary Figure S8

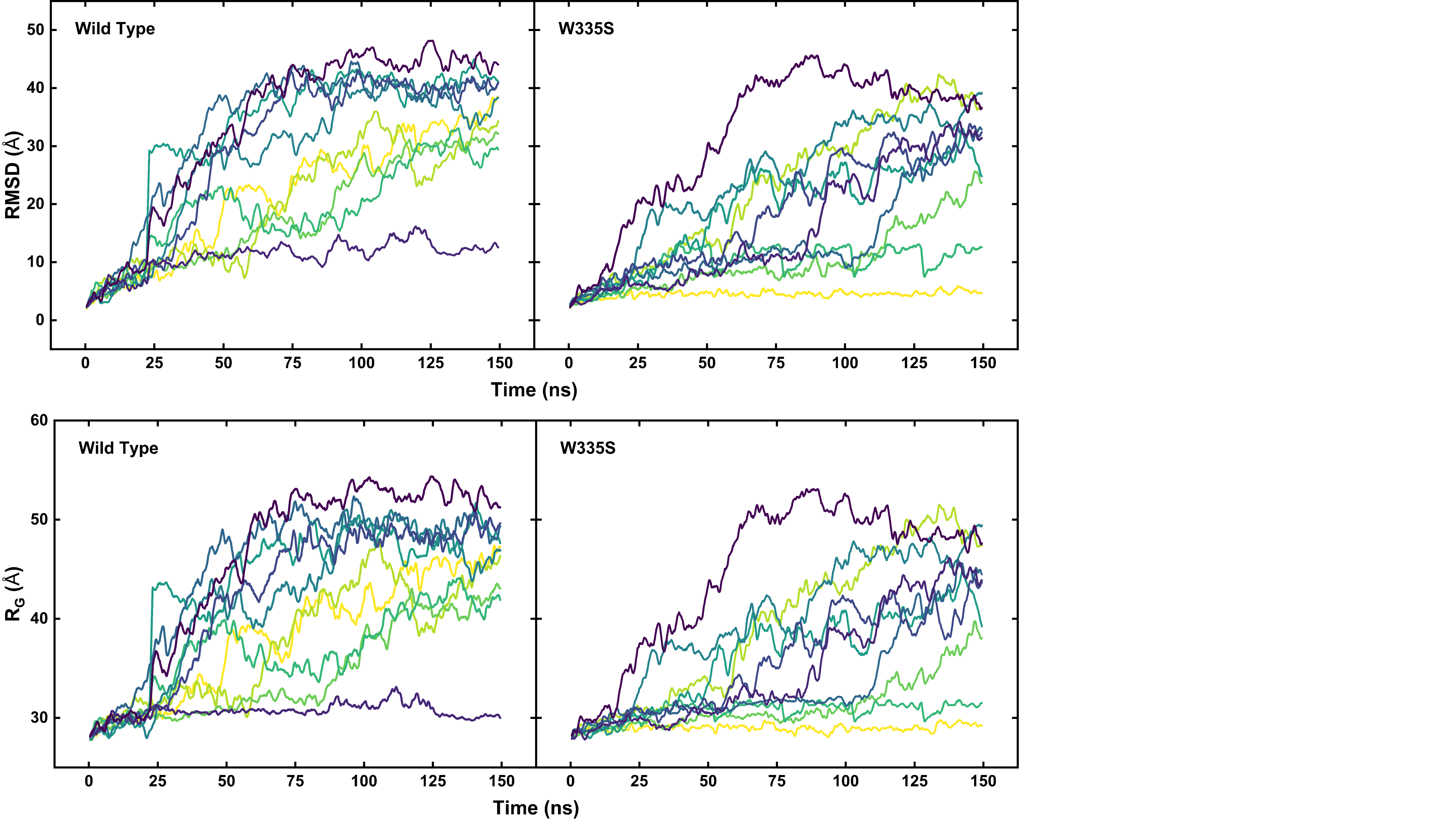

### Supplementary Figure S9

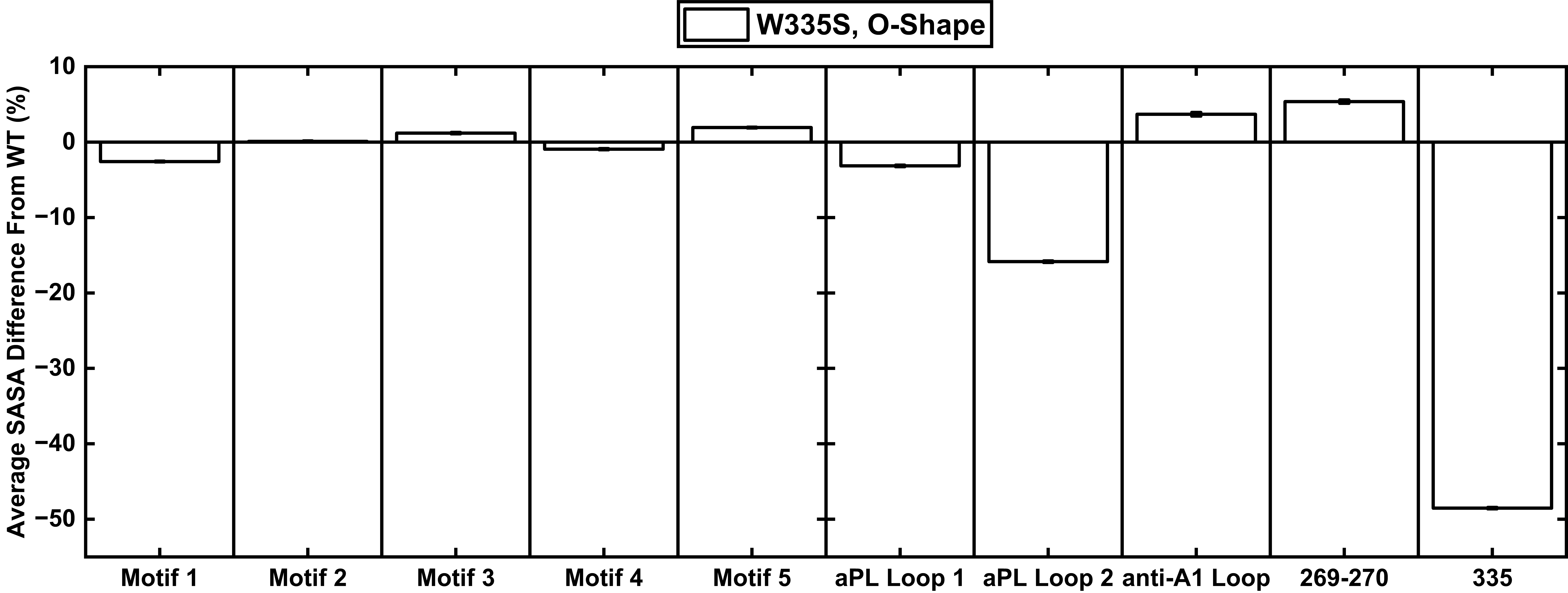
